# Optimizing the timing of one or two doses of pneumococcal conjugate vaccines in older adults in the United States: a modeling study

**DOI:** 10.1101/2025.10.24.25338736

**Authors:** Deus Thindwa, Anabelle Wong, Matthieu Domenech de Cellès

## Abstract

**Background:** Older adults have a high burden of pneumococcal disease. Pneumococcal conjugate vaccines (PCVs) are effective in reducing disease risk. Recent changes to lower recommended age of pneumococcal vaccination among PCV-naïve adults by the United States (US) Centers for Disease Control and Prevention (CDC) raises important questions on the optimal age of pneumococcal vaccination, as well as revaccination later in life when immunogenic responses to vaccination declines and disease activity peaks. We aimed to estimate the optimal age(s) and impact of a single dose and two-dose series of pneumococcal vaccination in adults ≥50 years-old (y) in the US.

**Methods and findings:** Age– and vaccine-serotype-specific IPD cases from the CDC’s Active Bacterial Core surveillance (ABCs) system among adults ≥50y in 2022 were combine with age and race demographic estimates, status of prior PCV use in adult population, vaccine effectiveness (VE), and waning VE dynamics in a cohort model of vaccine impact. Of the 12.8 million ABCs residents aged ≥50y, 52.4% were aged ≥65y. Black adults in age band 50-64y accounted for 60.2% of the total Black adult population compared to 51.8% of non-Black adults among the total non-Black adult population. A total of 1,557 IPD cases were reported across all ABCs sites; 349 cases in Black (N=1.6 million) and 1,208 cases in non-Black (N=11.2 million) adults. Cases caused by serotypes covered in higher-valency PCV20, PCV21 and PCV31, respectively, accounted for 52.7%, 74.9% and 87.9% of all IPD cases, and these proportions were similar between Black and non-Black adults. The optimal age of a single-dose higher-valency PCV was at 60y or 65y in overall population; earlier at 60y in Black adults compared to 65y in non-Black adults if initial VE was not influenced by age of vaccination. The optimal age shifted to 50y or 60y in overall population; earlier at 50y in Black adults compared to 60y in non-Black adults if initial VE was lower at higher age of vaccination. For maximum impact, a two-dose strategy showed that if the first dose is given at ≥65y, the second dose should be given at 5 years after initial dose, and if the first dose is given between 50-64y, the second dose should be given at 10 or 15 years after initial dose, irrespective of VE assumptions or race group.

**Conclusions:** Age of vaccination with a single dose of PCV that prevents maximum IPD cases is generally at 60y or 65y, and earlier among underserved populations, reflecting CDC’s new changes to lower age of vaccination. Optimal age of vaccination with a second dose of PCV is influenced by the timing of the first dose. Our results are driven by population age structure, age-specific IPD risk, VE dependency on age and waning VE uncertainty. These findings could help inform cost-effective planning and future age– and risk-based PCV recommendations.

**Author summary:** *Why was this study done?:*

- Disparities in pneumococcal disease burden are widespread among adults aged ≥50 years-old in the United States across demographic groups
- This study was conducted to explore the optimal age of vaccination with a single dose of higher-valency pneumococcal conjugate vaccine (PCV) in older adults
- This study was conducted to evaluate the potential benefits of a booster dose with a higher-valency pneumococcal conjugate vaccine and the optimal timing of that dose

*What did the researchers do and find?:*

- We conducted a modeling study using a cohort model and age– and serotype-specific invasive pneumococcal disease (IPD) case data from the Centers for Disease Control and Prevention (CDC)’s large Active Bacterial Core surveillance (ABCs) system, stratified by epidemiologic and vaccination variables
- The age of vaccination with a single dose of higher-valency PCV that averted most cases was overall at 60y or 65y (60y for Black adults, 65y for non-Black adults) for age-independent initial VE, or overall at 50y or 60y (50y for Black adults, 60y for non-Black adults) for age-dependent initial VE.
- Optimal timing of a second dose of higher-valency PCV was at 5 years after administration of initial dose in late adulthood (≥65y) or at 10 or 15 years after administration of initial dose in early adulthood (50-64y)

*What do these findings mean?:*

- Age of vaccination with a single dose of higher-valency PCV that averts most cases is earlier among Black than non-Black adults
- Appropriate timing of a second dose of PCV can improve the management of pneumococcal disease
- Studies are needed to provide evidence of long-term changes in PCV effectiveness against pneumococcal disease among older adults

## Introduction

*Streptococcus pneumoniae* (pneumococcus) causes a high burden of disease among older adults, including bacteremia and meningitis [1]. Pneumococcal conjugate vaccines (PCVs) are effective at reducing the burden of disease caused by vaccine serotypes [2–6]. In the United States (US), a 13-valent pneumococcal conjugate vaccine (PCV13) and a 23-valent pneumococcal polysaccharide vaccine (PPSV23) have been used to prevent pneumococcal disease in adults aged ≥65 years-old (y) [7,8]. More recently, PCV15, PCV20 and PCV21 have also been licensed and recommended to replace PCV13 [7,9–12]. Next generation vaccines targeting more than 30 serotypes are under development for use in adults and are progressing through clinical testing [13].

The use of PCVs in infants has reduced the transmission of vaccine-targeted serotypes (VTs) in the community and has consequently reduced the burden of disease caused by these serotypes in older adults [14–16]. However, a substantial burden of pneumococcal disease caused by non-vaccine serotypes (NVTs), persistent VTs and re-emerging VTs remains in older adults [17–22]. For instance, serotypes targeted by PCV13 still accounted for 27.5% of all invasive pneumococcal disease (IPD) in 2022 among adults aged ≥50y [23], more than 10 years after PCV13 had been introduced in children and was recommended for older adults. This persistent or re-emerging burden caused by VTs is caused by imperfect effectiveness against targeted serotypes and, likely, by waning of protection over time [23]. This current burden of NVTs and VTs underscore a need for more effective and higher serotype coverage vaccines that match the circulating serotypes and whose administration is well-timed to maximize protection.

In 2024, the US Centers for Disease Control and Prevention (CDC) changed the recommended age for universal receipt of PCV from ≥65y to ≥50y among all PCV-naïve adults [24,25]. This change in the recommendation was largely done to ensure adequate protection during key risk periods. In particular, the age distribution of cases among Black Americans aged ≥50y skews substantially towards less than 65y than the age distribution for other demographic groups, and the overall life expectancy is around 72 years compared to 80 years for other groups combined [26]. Thus, a recommendation based on age 65y and above would have missed many cases and might have contributed to widening disparities in protection.

There are tradeoffs, however, in lowering the age of vaccine receipt. While vaccinating sooner ensures that fewer cases are missed among younger adults, protection resulting from vaccination likely does not last the entire lifespan. The risk of severe pneumococcal disease increases sharply with age, so waning of immunity could leave adults who survive to older ages vulnerable during the period of highest risk if they are immunized too soon [27]. Despite several age– and risk-based guidelines for PCV vaccination among adults in Europe and the US [25,28], there is little evidence to support age timing for a single dose or booster dose of PCV. Moreover, with the recent advancements in pneumococcal policy and epidemiology in the US including age shift in vaccination recommendation to ≥50y, and recommendation of broader-valency vaccines, evidence gaps are apparent on whether additional doses later in life when disease burden peaks maybe necessary. Such gaps have opened new and critical questions which can be addressed through modeling.

In this study, we investigated age-based optimization for (a) a single dose of higher-valency PCV20, PCV21 and investigational PCV31 and (b) a two-dose series comprising PCV13 as the first dose and PCV20, PCV21 or PCV31 as the second dose in adults aged ≥50y in the US under several epidemiologic and vaccination scenarios and assumptions. We also explored the effect of differential life expectancy on optimal age of vaccination, using variations between race groups to highlight the issue.

## Methods

### Population demographic data

We obtained the 2022 annual population counts of adults ≥50y from the US Census Bureau (https://www2.census.gov/programs-surveys/popest/datasets) [26]. The 2022 data contain the population size in the counties in 10 States where CDC’s Active Bacteria Core surveillance (ABCs) program is implemented [5]. Annual population estimates by age were smoothed using a 5-year moving average to reduce the effects of stochasticity in downstream results (Fig S1A) [27,29].

### Invasive pneumococcal disease data

The ABCs is the CDC’s long-standing active, population-based surveillance system for invasive bacterial pathogens and has been running since 1995 [5]. The ABCs catchment area represents approximately 12.8 million people aged ≥50y. The 2022 data on IPD surveillance used in this analysis included age, serotype, and IPD case counts. The 2022 period was chosen because pediatric PCV13 and adult-PCV13/PPSV23 policies were stably in place, and PCV15, PCV20 and PCV21 were not yet in widespread use. Similarly, we only considered IPD data in a mature infant-PCV program, at least 10 years post-PCV13 introduction and before PCV15/PCV20 replaced PCV13 in 2022/2023, respectively [7]. Timelines of vaccination introductions among infants and older adults are detailed in the supplement (Text S1).

### Race– and age-specific IPD incidence

IPD case counts by serotype are made publicly available in broad age categories: 50-64y and 65+y. To disaggregate the 65+y category, we used the ABC’s overall age distribution of IPD cases in this population for 65-74y, 75-84y, 85+y [5], and multiplied these proportions by the IPD case counts for the broader 65+y category. Data on racial groups were inferred by multiplying age-specific IPD case counts by reported age-specific proportion of IPD among Black and non-Black adults [24]. IPD incidence was calculated by dividing the number of race-, age– and vaccine serotype group-specific IPD cases by the race– and age-specific population counts and multiplied by 100,000. To interpolate and extrapolate IPD incidence to single-year age, we fitted an exponential growth model to race-, age– and vaccine serotype group-specific IPD case counts using a non-linear least square function. Thus, *E*(*I*|*a*) = *ye*^βα^ where *E*(*I*|*a*) is the expected IPD incidence at age *a* where β is the growth rate and *y* is the incidence at age 0. Uncertainty around incidence was obtained by bootstrap sampling fitted parameter means and covariance matrix 1000 times and then summarized through 95% quantiles of samples.

### Pneumococcal conjugate vaccine impact model

*Model overview*. We adapted and expanded our previous cohort model to simulate the risk for IPD in adults ≥50y as a function of age, race, population demographics and IPD incidence, and compute the expected number of IPD cases in single-year age cohorts [27]. The following outcomes definitions were used,

– Vaccine impact of a single-dose PCV was mathematically a function of the expected number of IPD cases, vaccine effectiveness (VE), waning of vaccine-derived immunity and dose timing [27].
– Vaccine impact of the second dose in a two-dose series PCV program was mathematically a function of the remaining number of IPD cases after averting initial IPD cases with the first dose of PCV, vaccine effectiveness, waning of vaccine-derived immunity of the second dose given the waning VE levels of the first dose, and dose timing.
– Vaccine impact (vaccine preventable number of IPD cases) was the lifetime number of cases averted through vaccinating 100% of older adults at five-year interval age under different vaccination and epidemiologic scenarios below.
– The proportion of preventable IPD cases was defined as the number of IPD cases that can be prevented if vaccination is administered at a certain age cohort divided by the total number of IPD cases that would have occurred in a cohort of adults ≥50y in the absence of immunisation.
– Vaccine efficiency of the program in a single-dose strategy was calculated as the number of individuals needed to vaccinate to prevent an IPD case or the number of doses needed to administer to an age cohort to prevent an IPD case [30,31].
– Vaccine efficiency of the program in a two-dose strategy was calculated as the number of first and second doses needed to administer to prevent an IPD case.

Our baseline/main analyses were based on the overall, Black and non-Black demographics, PCV being in use in the older adult population, age-independent initial VE, and fast and slow waning VE informed by the assumptions used in a previous modeling exercise conducted for the CDC’s Advisory Committee on Immunisation Practices (ACIP). Time steps in the model were set at one year. Age-dependent all-cause mortality was set to match observed population demographics of older adults in the US, whereas age-dependent IPD risks were set to match fitted IPD incidence in each single-year age.

*Inferring serotype age distribution assuming no current PCV program*. For the baseline analysis, we used reported IPD data in the US population where PCV13/PPSV23 program among adult aged ≥65y has been on-going for several years. In a sensitivity analysis, we considered an alternative scenario assuming that adult PCV13/PPSV23 had not been in regular use. To do this, reported IPD cases were back-inflated by combining data on PCV13/PPSV23 vaccination coverage of 65%, observed number of IPD cases, and equally-weighted vaccine effectiveness between PPSV23 and PCV13 of 58% in adults aged ≥65y [7,32,33]. Thus, *C*_1_ = *C*_2_/(1 − *v* ∗ *VE*_α_) where *C*_1_ is the estimated IPD case counts assuming no adult PCV13/ PPSV23 program, *C*_2_ is the reported IPD case counts in the presence of adult PCV13/PPSV23 program, *v* is the adult PCV13/PPSV23 coverage in 2022, and *VE*_α_” is the age-adjusted vaccine effectiveness of adult PCV13/ PPSV23 (Fig S1B).

*Initial vaccine effectiveness (VE) and waning VE*. In a October 2024 report by the ACIP [25], the initial PCV effectiveness was considered constant for 5 years from the time since vaccination as evident from clinical trials and effectiveness studies [8,34,35]. To inform PCV cost effectiveness analyses in older adults, the ACIP assumed waning VE to zero for the next 10 years (fast waning) or 15 years (slow waning) after the initial 5-year stability [25]. Our model simulated a constant initial VE of 68% (38-84) for 5 years based on a PCV effectiveness study in the US adult population [8], and then adapted the ACIP assumptions of waning VE to zero after 10 or 15 years of initial VE for fast and slow waning scenarios [25]. A linear function that captures the initial VE and waning VE was used. Mean VE and its 95%CI estimates were initially converted to log relative risk (log_*RR* = *log*(1 − *VE*/100)) to ensure *RR* is normally distributed before bootstrap sampling from a normal distribution centered on the mean log_*RR*_t_ at the time *t* with standard deviation (*sd*) derived from 95% Interval of log_*RR* as (*sd* = (log_*RR_max_* − log_*RR_min_*)/4). Then log_*RR* were converted back to VE (*VE* = 100 ∗ (1 − *exp*(log_*RR*))) for use in downstream analysis. The VE dynamics of a second dose in a two-dose series PCV scenario was a recursion of a single dose from the time of its administration. For instance, if the first dose is given at 50y, then (a) the second dose given at 55y would imply extending a 5-year constant VE of 68% for another 5 years to 10 years before waning resumes, or (b) the second dose given at 65y would imply boosting the initial dose from mean VE of 0% (under fast waning scenario) or 28% (under slow waning scenario) back to 68% for the next 5 years before waning resumes (Fig 1, Fig S4). In a sensitivity analysis, we fitted a stepwise decay model based on different assumptions of waning VE [27], to compare how its subsequent optimal age of vaccination would differ from the ACIP assumption of waning VE (Text S2)

**Figure 1.**
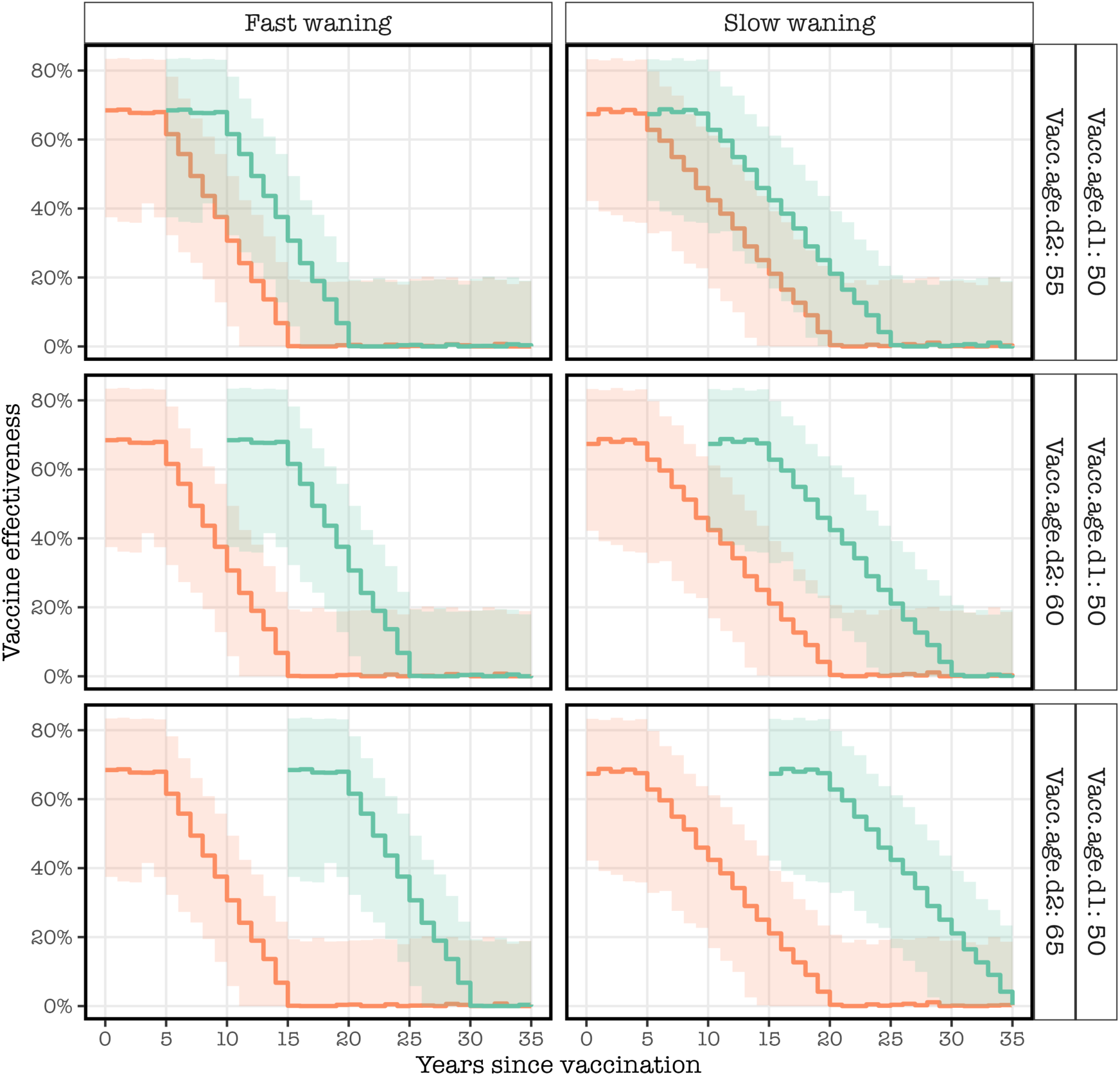
A snapshot of initial vaccine effectiveness (VE) and waning VE from the time since vaccination based on Advisory Committee on Immunisation Practices (ACIP) assumptions, stratified by age of vaccination of the first and second doses, and waning VE category under the age-independent initial VE. The blue solid line and shaded ribbon refer to mean VE and 95% confidence intervals (CI) of the mean VE of the first dose whereas the red solid line and shaded ribbon refer to mean VE and 95% CI of the mean VE of the second dose. The plot contrasts between fast and slow waning VE and subsequent 5-year age intervals of second dose vaccination for a given first dose at 50y.

*Age-dependency of initial VE*. There is no clear evidence of differential initial VE of PCV by age at administration. However, point estimates of initial VE in a PPSV23 effectiveness study in England showed declining initial VE with age [33], suggesting partially impaired immune responses to vaccination in late older adults. Thus, we explored a scenario of age-dependent initial VE of PCV in a sensitivity analysis. As in previous study [33], we assumed that VE would be 22%, 33% and 44% less of initial VE if PCV is given at ≥55y, ≥65y and ≥75y, respectively.

*Timing of second dose of higher-valency PCV after first dose of PCV13.* Vaccines targeting different serotypes include PCV13 (1, 3, 4, 5, 6A, 6B, 7F, 9V, 14, 18C, 19A, 19F, 23F), PCV20 (1, 3, 4, 5, 6A, 6B, 7F, 8, 9V, 10A, 11A, 12F, 14, 15B, 18C, 19A, 19F, 22F, 23F, 33F), PCV21 (3, 6A, 7F, 8, 9N, 10A, 11A, 12F, 15A, 15B, 16F, 17F, 19A, 20A, 22F, 23A, 23B, 24F, 31, 33F, 35B), and investigational PCV31 (1, 2, 3, 4, 5, 6A/C, 6B, 7F, 7C, 8, 9N, 9V, 10A, 11A, 12F, 14, 15A, 15B, 16F, 17F, 18C, 19F, 19A, 20B, 22F, 23F, 23A, 23B, 31, 33F, 35B). We explored the optimal age and impact dynamics of vaccinating with a single dose using PCV20, PCV21, or PCV31 following a single dose of PCV13 to reflect the status quo where the majority of US older adults (≥65y) that were vaccinated with PCV13 may need a second dose of higher-valency PCV. For a given age of vaccination of the first dose of PCV13, we explore all possible 5-year interval age of vaccination of second dose of higher-valency PCV. We also show alternative high-resolution age of vaccination intervals using 2 years as an example.

Statistical significance was considered at *p*<0.05. All the models and statistical analyses were respectively developed and conducted using R language v4.1.1 [36]. The R code and data are publicly accessible and archived on Github [37].

### Ethical approval

This study did not require any ethical approval. All the datasets used in this study were obtained from publicly available repositories and documents. Individual patient informed consent was not required for the use of publicly available anonymised and aggregated routine surveillance data.

## Results

### United States population demographics and IPD incidence

Among the 12.8 million US adults aged ≥50y in 2022, 47.2% were aged ≥65y and 5.2% were aged ≥85y. The population age distribution varied by race, with 60.2% of the ≥50y Black population in the 50-64y age band compared to 51.8% of the non-Black population. A total of 349 and 1,208 serotype-specific IPD cases among Black (N=1.6 million) and non-Black (N=11.2 million) adults were reported in 2022 across all ABCs sites. Of all IPD cases, cases caused by serotypes covered in PCV20, PCV21 and PCV31 accounted for 52.7%, 74.9% and 87.9%, respectively, and these proportions were similar between Black and non-Black population (Fig 2, Fig S2).

**Figure 2.**
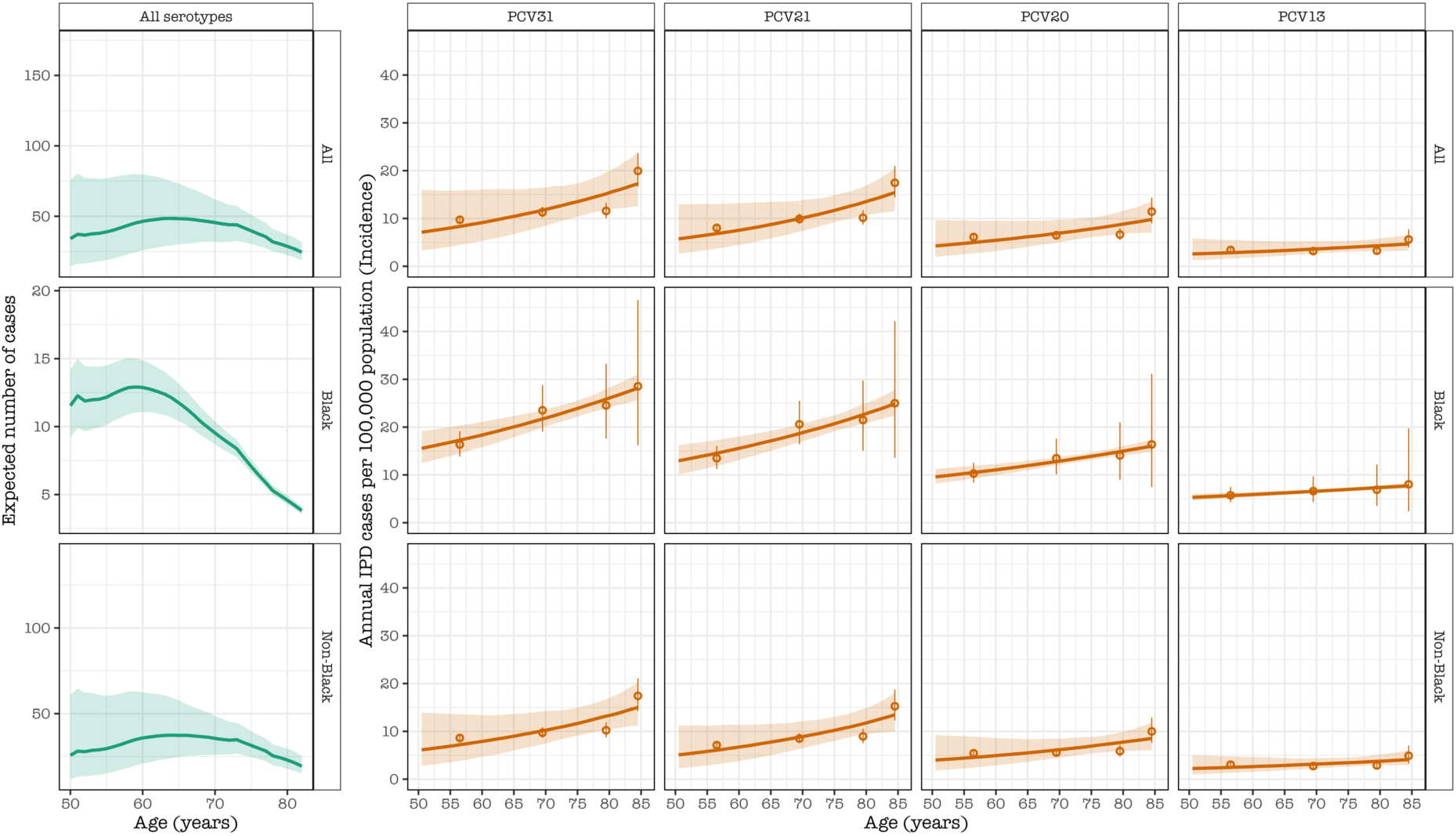
The expected number of invasive pneumococcal disease (IPD) cases, caused by all serotypes, peak in the early-age of Black adulthood and in the middleage of non-Black adulthood (green). The estimated incidence of IPD by demographic groups and serotype group covered in each PCV product in single-year age in the presence of prior PCV immunity among the adult population in the United States (orange). The green and orange solid line and shaded ribbon refer to the mean expected number of cases or incidence and their respective 95% confidence intervals (CI) of the mean. The orange circles and vertical lines refer to the inferred or reported IPD cases per 100,000 population and corresponding 95%CI.

Overall IPD incidence increased sharply by age, but the difference in incidence by age between 85y vs 50y was more pronounced for the non-Black population (incidence risk ratio [IRR]: 2.11, 95% Confidence Intervals [95%CI]: 1.95-2.27) than the Black population (IRR: 1.63, 95%CI: 1.25-2.02). Because the age distribution is skewed younger in the Black population, a majority of IPD cases occurred in the 50-64y population, while for the non-Black population, most cases occurred among people aged ≥60y (Fig 2, Fig S2, Fig S3).

### Optimal age for vaccination with a single dose of higher-valency PCV

The optimal age of immunisation depends on the age distribution of cases and the life expectancy of the population and differs for the Black and non-Black individuals. Based on the current data from the US (where there has already been substantial PCV13 use), and regardless of the assumptions of waning VE, the optimal age of vaccination is 60y or 65y for overall population; 60y among Black adults and 65y among non-Black adults. The highest number of cases averted by each higher-valency PCV is consistently achieved under slow waning VE compared to fast waning VE. The proportions of prevented IPD by each PCV were steadily higher among Black than non-Black adults and higher for slow than fast waning VE (Figure 3, Table S1).

**Figure 3.**
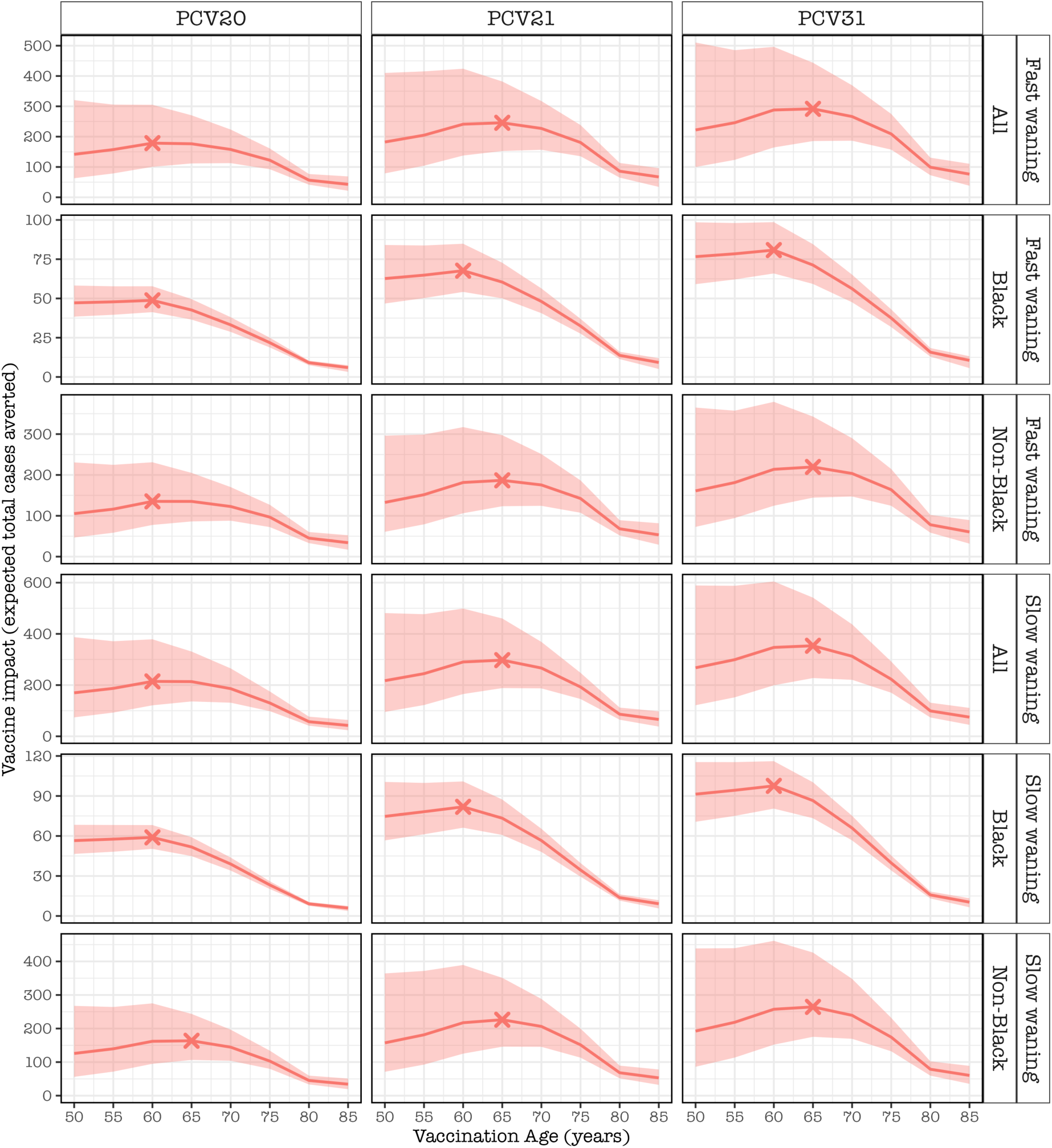
The expected number of IPD cases averted for the rest of the age cohort lifetime by vaccinating every older adult in the age cohort, stratified by pneumococcal conjugate vaccine (PCV) product, demographic group and waning PCV effectiveness (VE) category, under the scenarios of ACIP age-independent initial VE and waning VE and presence of prior PCV immunity in the adult population in the United States. The red line and shaded ribbon represent the cohort model mean estimates and 95% bootstrap confidence intervals for the mean estimates. The X corresponds to the optimal age for pneumococcal vaccination. Among Black adults, most cases are preventable at 60 years whereas among non-Black or overall adult population, this is achieved at 60y or 65y.

### Vaccination efficiency under a single-dose strategy of PCV

The lowest number of older adults that need to be vaccinated with a single dose of a PCV to prevent a case of IPD (vaccination efficiency) is achieved at an earlier age among Black adults compared to non-Black adults or overall adult population under the fast-waning VE scenario. In contrast, vaccination efficiency is achieved at the same age for the two groups under the slow-waning VE scenario. Under fast-waning VE, the optimal age of vaccination efficiency is 70y among Black adults and 75y among non-Black adults or overall adult population, irrespective of PCV product, whereas under slow waning VE, the optimal age of vaccination efficiency is 70y irrespective of race demographics. Thus, 520 (320-660) and 1,060 (660-1,640) of PCV31 Black and non-Black vaccinees, 600 (370-780) and 1,250 (770-1,960) of PCV21 vaccinees, and 880 (560-1110) and 1,780 (1100-2,750) of PCV20 vaccinees aged 70y are needed to optimally prevent a case of IPD under the slow waning VE scenario (Fig 4, Table S2).

**Figure 4.**
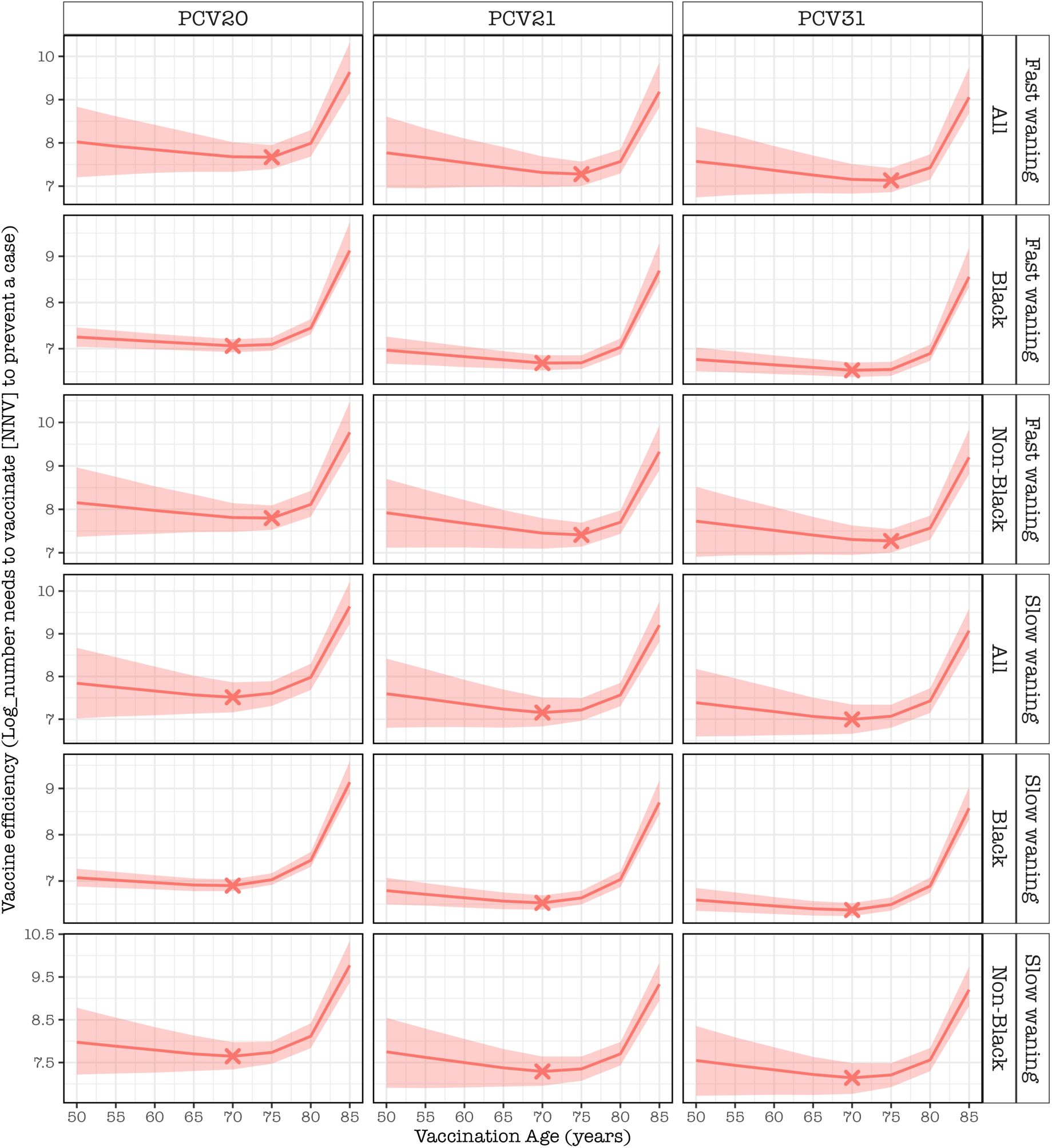
The logarithm of number needed to vaccinate (NNV) to prevent a case of invasive pneumococcal disease (IPD) in each age of vaccination, stratified by pneumococcal conjugate vaccine (PCV) product, demographic group and waning PCV effectiveness (VE) category under the scenarios of ACIP initial and waning VE, age-independent initial VE and presence of prior PCV immunity in adult population in the United States. The red lines represent cohort model mean estimates, and the shaded red ribbon represents 95% bootstrap confidence intervals for the mean estimates. The X represents the optimal age for vaccine efficiency. The optimal age of vaccine efficiency occurs at 70y for slow waning VE irrespective of race demographics whereas for fast waning VE it occurs at 70y among Black adults or 75y among non-Black adults and overall adult population.

### Optimal ages for vaccination with two-dose series of PCVs

We evaluated the optimal combination of the age of the first dose of PCV13 and age of a second dose of PCV20, PCV21 or PCV31. The optimal ages, at which the maximum number of IPD cases could be prevented, is achieved under two general situations regardless of waning VE scenario, PCV product or racial group; (a) if the first dose of PCV13 is given at ≥65y, then the second dose of higher-valency PCV should be given at 5 years of initial dose, or (b) if the first dose of PCV13 is given between 50-64y, then the second dose of higher-valency PCV should be given at 10 or 15 years after the initial dose. We show this with an example of introducing PCV31 as a second dose following the first dose of PCV13 under slow waning VE and age-independent initial VE in the overall, Black and non-Black populations, with other scenarios shown in the supplementary (Fig 5, Fig S5).

**Figure 5.**
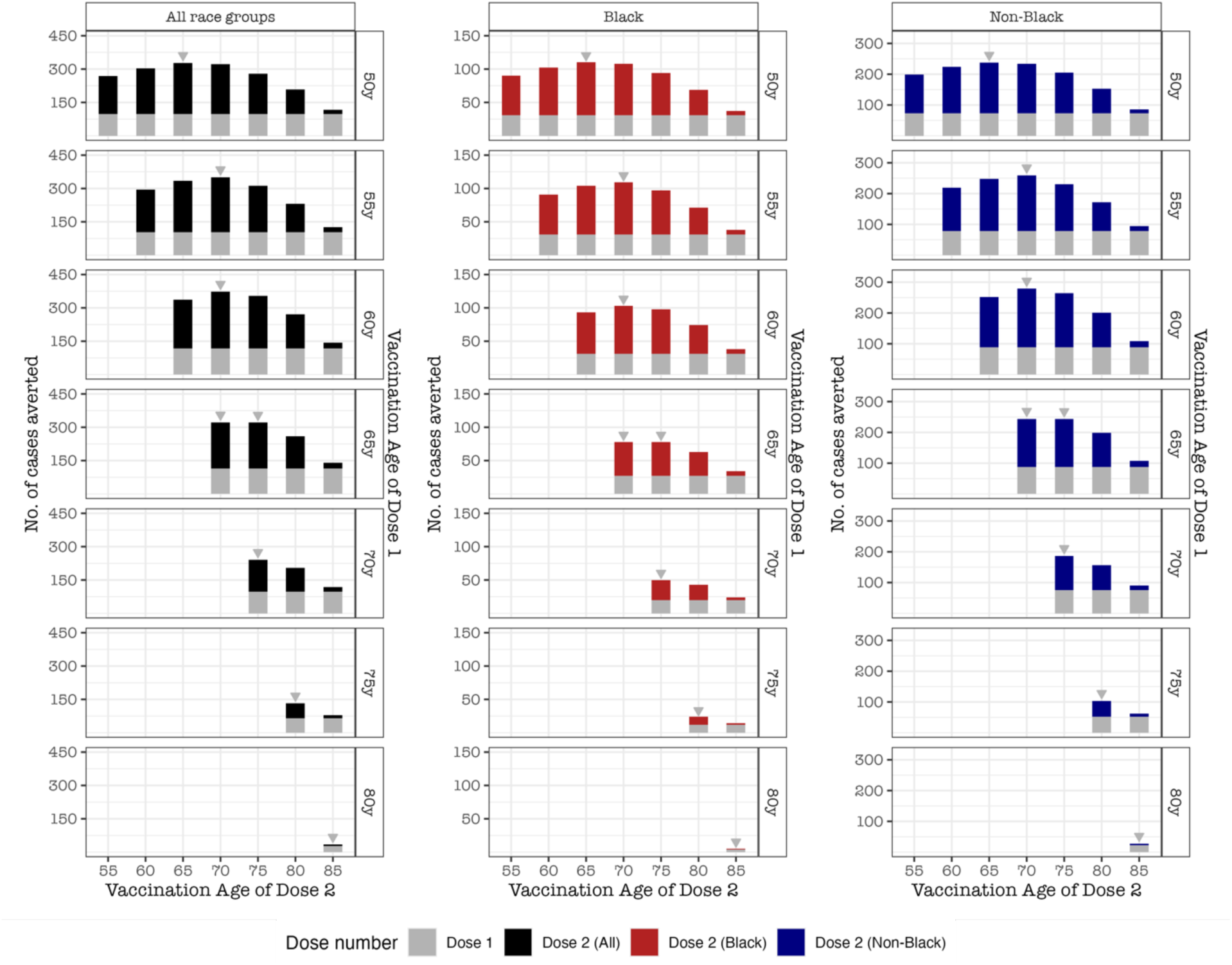
The predicted number of IPD cases averted for the rest of the age cohort lifetime for administering the first dose of PCV13 at a specific age (right y-axis) followed by the second dose of higher-valency PCV at any of the ages (x-axis), stratified by pneumococcal conjugate vaccine (PCV) product, demographic group and waning PCV effectiveness (VE) category, under the scenarios of ACIP initial and waning VE and presence of prior PCV immunity in adult population in the United States. The bars show cohort model mean estimates and the red color indicate second dose Black vaccinees and blue color indicate non-Black vaccinees. The optimal ages of the first and second doses are similar regardless of demographics. The second dose should be given at 5 years later if receipt of first dose was in late adulthood (65y+) or the second dose should be given at 10 or 15 years later if receipt of first dose was in early adulthood.

**Table.**
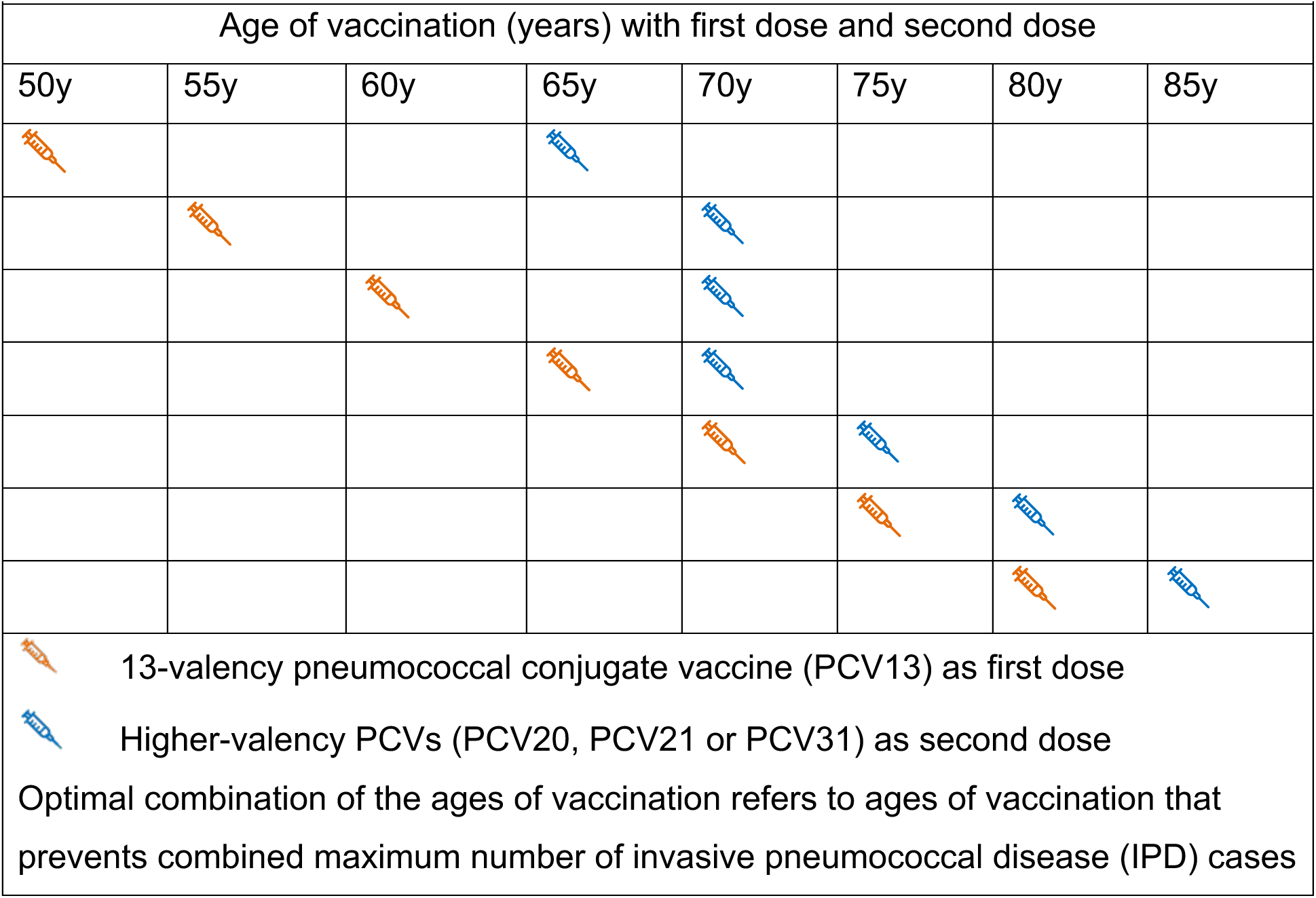
Summary of the recommended optimal combination of doses at different ages of vaccination in years (y) using the first dose of PCV13 and second dose of a higher-valency pneumococcal conjugate vaccine (PCV)

Our model specifically predicts that in order to avert the maximum number of IPD cases; the first dose of PCV13 given at 50y or 55y should be followed by a second dose of higher-valency PCV at 15 years later, the first dose of PCV13 given at 60y should be followed by a second dose of higher-valency PCV at 10 years later, and the first dose of PCV13 given at 65y, 70y, 75y or 80y should be followed by a second dose of higher-valency PCV at 5 years later (Table).

### Vaccination efficiency under a two-dose strategy of PCV

The total number of first and second doses needed to prevent a case of IPD (vaccination efficiency) is minimum (optimal) when a PCV31 second dose is administered at age of 80y irrespective of the age of receipt of the first dose of PCV13 in the overall population or among each race group. For instance, to optimally prevent a case of IPD, the number of first and second doses needed would total to 770 (408-1,297) at the age of 50y and 80y, 700 (407-1,099) at 55y and 80y, 609 (397-885) at 60y and 80y, 635 (452-852) at 65y and 80y,and 810 (624-1,056) at 70y and 80y (Fig 6 Table S3).

**Figure 6.**
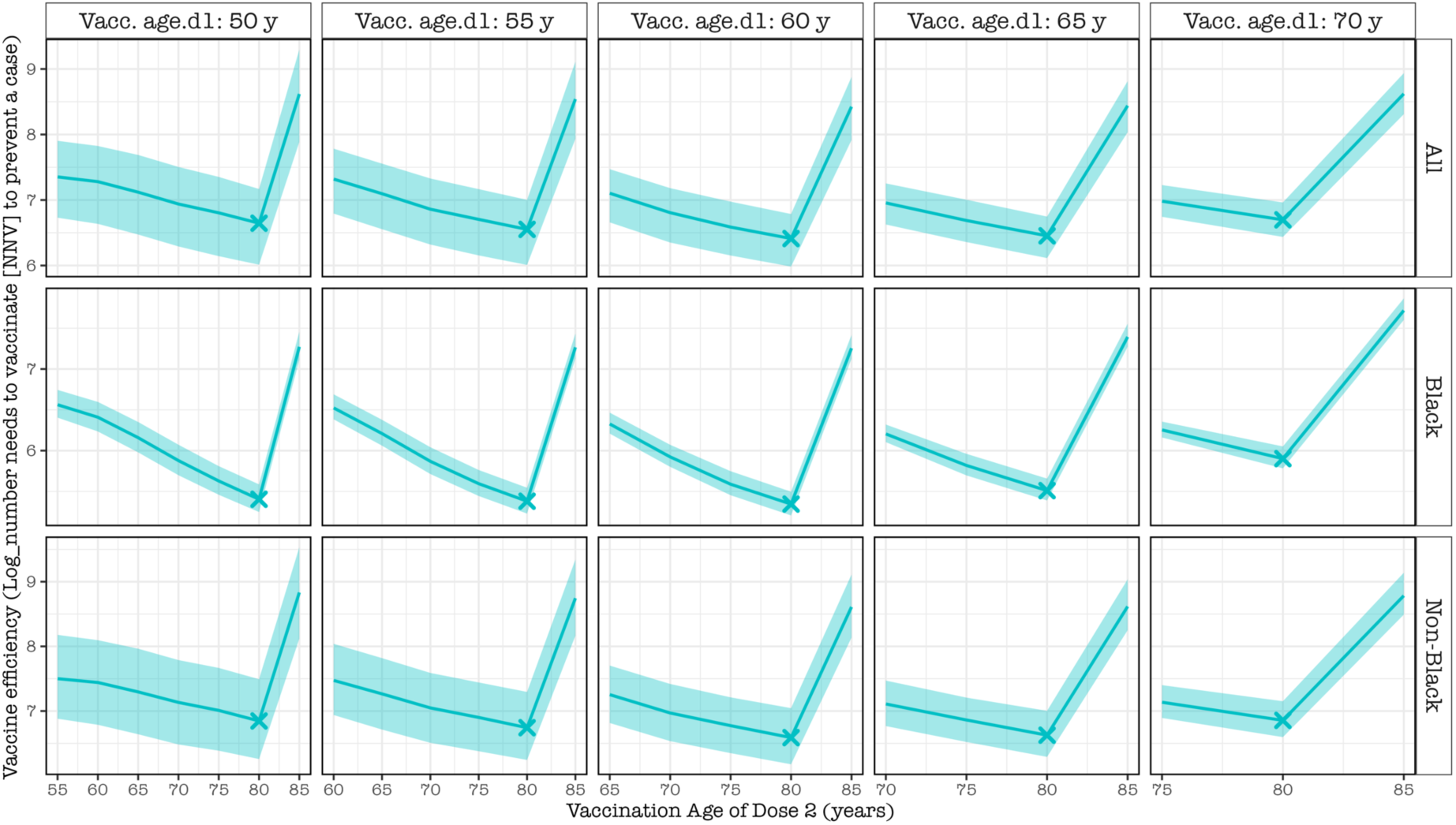
The logarithm of number needed to vaccinate (NNV) to prevent a case of invasive pneumococcal disease (IPD) in each age of vaccination of second dose PCV31, stratified by age of vaccination of the first dose PCV13, demographic group under the scenarios of ACIP initial and slow waning VE, age-independent initial VE and presence of prior PCV immunity in the adult population in the United States. The blue lines represent cohort model mean estimates, and the shaded blue ribbon represents 95% bootstrap confidence intervals for the mean estimates. The X represents the optimal age for vaccine efficiency. The optimal age of second dose vaccine efficiency occurs at 80y among all race demographics irrespective of the age of receipt of first dose of PCV13.

### Results from sensitivity analyses

Sensitivity analyses that evaluated the impact of different modeling assumptions, some of the specific results changed, but the overall conclusions remain the same. The most influential factor affecting optimal age timing was whether the initial VE varied with age. The functional form of the waning (i.e., step change vs ACIP assumption of linear waning) was not influential. The specific effects of changing these assumptions varied by risk group. If initial VE was age-dependent, there was a left shift in optimal age of vaccination e.g., the optimal age of vaccination for Black adults shifted to 50y from 60y (baseline scenario) whereas for non-Black adults shifted to 60y from 65y (baseline scenario) (Fig S6). If prior PCV immunity was absent in the adult population (‘PCV naïve adult population’) under the age-independent initial VE, the optimal age of vaccination generally reflected the baseline scenario at 65y among non-Black adults and 60y among Black adults (Fig S7). Likewise, assuming PCV naïve adult population under the age-dependent initial VE shifted the optimal age to 50y among Black adults and to 60y among non-Black adults (Fig S8). If stepwise decay of waning VE was assumed (Fig S9); under the presence of prior PCV immunity in adult population and age-independent initial VE, then the optimal age of vaccination is overall 60y (50y among Black adults and 60y among non-Black adults), otherwise under the age-dependent initial VE, the optimal age of vaccination is 50y irrespective of demographics (Fig S10). Lastly, in the absence of prior PCV immunity in adult population, optimal age of vaccination under the age-dependent or independent initial VE is, respectively, 50y or 60y among Black adults and 60y or 65y among non-Black adults (Fig S11).

## Discussion

We have estimated the optimal age for pneumococcal vaccination among older adults in the US using two strategies: (a) a single dose of higher-valency PCV20, PCV21, or PCV31 and (b) a two-dose series comprising PCV13 as the initial dose followed by a higher-valency PCV as the second dose. Our results show that the optimal age for a single dose of a higher-valency PCV in the overall population is approximately 60y or 65y under age-independent initial VE and 60y or 50y under age-dependent initial VE. For a two-dose strategy, when the first dose is administered at ≥65y, the second dose should ideally be given at 5 years later or when the first dose is given between 50-64y, the second dose should be given later at 10 to 15 years, irrespective of waning VE or racial subgroup. These findings are primarily driven by population age structure, age-specific IPD risk, age dependency of initial VE, and uncertainty surrounding waning immunity. In light of the recent CDC policy shift to vaccinate PCV-naïve adults aged ≥50y (rather than ≥65y), and the growing importance of revaccination strategies due to waning protection, our model provides timely and essential evidence to inform the optimal timing of both single-dose and two-dose PCV vaccination strategies.

Our model-derived estimates of the optimal age of vaccination were informed by ACIP assumptions regarding the waning of PCV effectiveness beyond five years (e.g., declining to 0% after 10 or 15 years) [24]. Although waning VE of PCVs among older adults likely affects the optimal timing of vaccination, empirical evidence beyond five years post-vaccination remains limited and warrants further investigation [8,34]. Notably, when alternative waning assumptions were applied using a stepwise decay model [27], where vaccine effectiveness does not decline to zero, the estimated optimal ages of vaccination were largely consistent with those generated under the ACIP-based assumptions (Fig S9, Fig S10, Fig S11).

The estimated optimal age for administering a single dose of higher-valency PCV was earlier among Black adults (at 60y or 50y) compared with non-Black adults (at 65y or 60y), respectively, regardless of assumptions about waning VE. This finding suggests that the optimal vaccination age may vary across subpopulations, likely reflecting differences in life expectancy and age-specific IPD risk [24,26]. Our results indicate that earlier vaccination among Black adults yields greater overall impact; however, this prediction should be interpreted in the broader context of other high-risk populations. From a policy perspective, any subgroup-specific recommendations must be balanced against practical considerations and the potential challenges introduced by increased programmatic complexity.

Evidence regarding the optimal timing for multi-dose administration of PCVs among older adults remains limited [28]. Although our modeling framework is capable of assessing all possible combinations of PCVs in a two-dose strategy, we focus here on a specific scenario in which the first dose is PCV13 and the subsequent dose is any considered higher-valency PCV. Our results suggest that the optimal timing of the second higher-valency PCV dose, whether administered earlier (when the first dose is given in late adulthood) or later (when the first dose is given in early adulthood), is primarily influenced by the interaction between age-specific IPD risk and the age distribution of the adult population eligible for vaccination. Administering the second dose earlier may be warranted when the elevated IPD risk in late adulthood outweighs the advantage of vaccinating a larger pool of younger adults, whereas delayed administration may be preferable when this balance is reversed.

From a practical standpoint, these findings suggest that, to maximize the public health impact of PCV use, adults aged ≥65y who recently received PCV13 under the US adult PCV program may benefit from receiving a higher-valency PCV approximately five years after their initial dose. Our primary analyses were based on five-year age of vaccination intervals and should thus be interprented with caution – a high-resolution assessment (e.g., using two-year intervals of age of vaccination) further refines this recommendation, indicating that individuals who received their first PCV13 dose in late adulthood (e.g., ≥70y) would likely benefit from receiving the second dose as soon as possible (Fig. S12).

Pediatric PCV programs have conferred substantial indirect protection against pneumococcal disease among older adults [2,14,38]. Our model did not explicitly account for this indirect (herd) protection, as it is likely to be non-differential across older adult age cohorts and therefore unlikely to influence the estimated optimal age of vaccination [39]. However, if children under-five years of age have distinct social contact patterns with grandparents (typically aged ≥65y) compared with adults aged 50-64y, the optimal vaccination age could shift, and the vaccine’s overall impact might be underestimated. In addition, our analysis stratified the US population only by Black and non-Black subgroups due to insufficient data to support finer demographic stratification. Consequently, the estimated optimal age of vaccination for these groups should be interpreted as broadly representative of underserved and socially advantaged populations, respectively. Although comorbidities are a key component of the CDC’s risk-based PCV recommendations, we did not model optimal age of vaccination by comorbidity status because of the absence of publicly available data.

Our model builds upon a previously established cohort modeling framework [27], extending its methods to incorporate ACIP assumptions regarding waning VE and to capture the dynamic interaction between the first dose and second doses of higher-valency PCVs. We utilized extensive age– and serotype-specific IPD data from the CDC’s ABCs system, a nationally representative US dataset, to inform the optimal age for administering a single dose or a two-dose series of PCV [5]. This study addresses key policy gaps by providing evidence on the vaccination ages and dosing intervals that could avert the greatest number of IPD cases among US adults. Beyond supporting the CDC’s recent recommendation to lower the age threshold for single-dose PCV administration to ≥50y [24], our findings suggest that optimizing the timing of two-dose PCV schedules across demographic groups could help to further enhance pneumococcal disease prevention. Future research should also explore precision vaccination strategies that tailor PCV selection and scheduling to community-level epidemiological patterns including local IPD serotype distributions (census tract) to further improve pneumococcal disease control in the US.

In conclusion, the optimal age of vaccination with a single dose of higher-valency PCV to prevent the greatest number of IPD cases is approximately 60y or 65y in the overall adult population, with an earlier optimal age predicted among Black adults compared with non-Black adults. The optimal timing of a second dose of higher-valency PCV is estimated at 5 years after the initial dose in late adulthood, and at 10 to 15 years after the initial dose in early adulthood. Overall, our findings provide evidence to support cost-effectiveness analyses and to guide future age– and risk-based PCV vaccination policies.

## Data availability

The aggregated and anonymised full datasets used in this study are freely available in the GitHub repository [37]

## Code availability

An R script used to analyze aggregated and anonymised datasets are freely available in the GitHub repository [37]

## Acknowledgements

This work was supported by Vaxcyte. The funder had no role in the study design; collection, analysis, or interpretation of data; manuscript writing and preparation; or decision to submit for publication. We would like to thank Professor Daniel M Weinberger of Yale University for the insightful discussion throughout this work and his contribution to editing the manuscript.

## Author contributions

Deus Thindwa: Writing – original draft, Writing – review & editing, Methodology, Investigation, Formal analysis, Data curation, Conceptualization. Anabelle Wong: Writing – original draft, Writing – review & editing, Methodology, Investigation, Formal analysis, Data curation, Conceptualization. Matthieu Domenech de Celles Writing – original draft, Writing – review & editing, Methodology, Investigation, Formal analysis, Data curation, Conceptualization. All authors read and approved the final manuscript.

