## Supplementary_file for "Optimizing the timing of one or two doses of pneumococcal conjugate vaccines in older adults in the United States: a modeling study"

**Title**

Deus Thindwa

**Table of contents**


| | |
|---|---|
| Text S1 | Description of Infant and older adult vaccination timelines |
| Text S2 | Description of stepwise decay model of vaccine effectiveness dynamics |
| Table S1 | Proportion of preventable IPD with a single dose of each vaccine |
| Table S2 | Number needed to vaccine with single dose to prevent a case of IPD |
| Table S3 | Number needed to vaccine with a second dose to prevent a case of IPD |
| Figure S1 | Population age distribution and number of IPD cases by racial groups |
| Figure S2 | Number of IPD cases by racial and vaccine serotype groups |
| Figure S3 | Inferred incidence of IPD by racial and vaccine serotype groups |
| Figure S4 | Snapshot of ACIP initial VE & waning VE from time since vaccination |
| Figure S5 | Combined impact of the first dose using PCV13 and second dose of higher valency PCV |
| Figure S6 | Vaccine impact assuming ACIP age-dependent initial VE and waning VE and presence of prior PCV immunity |
| Figure S7 | Vaccine impact assuming ACIP age-independent initial VE and waning VE and absence of prior PCV immunity |
| Figure S8 | Vaccine impact assuming ACIP age-dependent initial VE and waning VE and absence of prior PCV immunity |
| Figure S9 | Snapshot based on stepwise decay model of initial VE & waning VE from time since vaccination |
| Figure S10 | Vaccine impact assuming the scenarios of stepwise initial VE and waning VE and presence of prior PCV immunity |
| Figure S11 | Vaccine impact assuming the scenarios of stepwise initial VE and waning VE and absence of prior PCV immunity |
| Figure S12 | Combined impact of the first dose using PCV13 and second dose of higher valency PCV31 under high-resolution vaccination age intervals of 2-years |

**Suplementary Text 1**

*Infant vaccination timelines.* In July 2000, routine infant-PCV7 was introduced under a schedule of three primary doses at 2, 4, 6 months and a booster at 12–15 months (3p+1) [1,2]. Routine coverage increased rapidly within the first couple of years after introduction [1]. Later, PCV13 was licensed and replaced PCV7 in 2010 [3], and then PCV15 and PCV20 were also licensed and replaced PCV13 in 2022 and 2023 [4].

*Older adult vaccination timelines*. Among older adults aged ≥65y, a PPSV23 has been in use since the 1980s to prevent pneumococcal disease, and vaccination coverage slowly increased to current levels of ~65% [4]. In 2014, PPSV23 was complemented with PCV13 to improve effectiveness and duration of protection [4–6]. In 2021-2024, PCV15, PCV20 and PCV21 were also licensed and recommended to replace PCV13 [3,7,8]. Several other next-generation PCVs that target more than 20 serotypes are in clinical trials [9–11]. More recently, in October 2024, the United States Centers for Disease Control and Prevention (CDC)'s Advisory Committee on Immunization Practices (ACIP) lowered the age of PCV vaccination among PCV naïve older adults from ≥65y to ≥50y to cover a more at-risk population [12].

**Suplementary Text 2**

*Stepwise decay model*. Based on a systematic review [13], we adapted evidence that PCV would remain stable at 68% for 5 years [14], and then decline similar to two effectiveness studies of PPSV23 in the United Kingdom [15,16]. We modelled VE dynamics as a function of time since vaccination using piecewise constant models (Fig S9) [17]. We considered waning VE from Andrews et al. as fast waning VE [15], and from Djennad et al. as slow waning VE [16]. The VE was sampled using bootstrap sampling from a normal distribution centred on the mean VE at that time and standard deviation derived from the reported 95% confidence interval.

| Supplementary Table 1. Proportion of preventable IPD with a single dose of each vaccine | | | | | | | | |
|---|---|---|---|---|---|---|---|---|
| | | | **Fast Waning VE** | | | **Slow waning VE** | | |
| **Vaccine** | **Vac.age** | **Race group** | **2.50%** | **50%** | **97.50%** | **2.50%** | **50%** | **97.50%** |
| PCV20 | 50 | Black | 20.3 | 25.2 | 30.8 | 24.8 | 30.1 | 36.1 |
| PCV20 | 55 | Black | 21.2 | 25.5 | 30.5 | 25.8 | 30.6 | 35.9 |
| PCV20 | 60 | Black | 22.0 | 26.0 | 30.8 | 26.9 | 31.3 | 36.3 |
| PCV20 | 65 | Black | 19.3 | 22.7 | 26.3 | 23.8 | 27.5 | 31.5 |
| PCV20 | 70 | Black | 15.2 | 17.6 | 20.2 | 18.0 | 20.7 | 23.4 |
| PCV20 | 75 | Black | 9.9 | 11.6 | 13.3 | 10.7 | 12.3 | 14.0 |
| PCV20 | 80 | Black | 3.9 | 4.8 | 5.6 | 4.0 | 4.8 | 5.6 |
| PCV20 | 85 | Black | 1.7 | 3.2 | 4.0 | 2.0 | 3.1 | 4.0 |
| PCV21 | 50 | Black | 25.6 | 33.3 | 43.1 | 31.0 | 39.8 | 51.1 |
| PCV21 | 55 | Black | 27.3 | 34.5 | 43.4 | 33.4 | 41.5 | 51.6 |
| PCV21 | 60 | Black | 29.4 | 36.1 | 43.7 | 36.0 | 43.4 | 51.9 |
| PCV21 | 65 | Black | 27.2 | 32.2 | 37.6 | 33.2 | 38.9 | 45.4 |
| PCV21 | 70 | Black | 21.8 | 25.6 | 29.2 | 26.2 | 30.0 | 34.3 |
| PCV21 | 75 | Black | 14.9 | 17.2 | 19.6 | 16.0 | 18.3 | 21.0 |
| PCV21 | 80 | Black | 6.1 | 7.3 | 8.5 | 6.1 | 7.3 | 8.6 |
| PCV21 | 85 | Black | 2.7 | 4.9 | 6.3 | 3.1 | 4.9 | 6.3 |
| PCV31 | 50 | Black | 32.3 | 40.7 | 50.9 | 38.7 | 48.6 | 59.1 |
| PCV31 | 55 | Black | 33.8 | 41.8 | 50.5 | 41.1 | 50.2 | 59.2 |
| PCV31 | 60 | Black | 35.7 | 42.9 | 51.0 | 43.7 | 51.9 | 60.0 |
| PCV31 | 65 | Black | 32.3 | 37.9 | 43.9 | 39.2 | 46.1 | 52.3 |
| PCV31 | 70 | Black | 25.8 | 29.9 | 34.1 | 30.3 | 35.1 | 39.6 |
| PCV31 | 75 | Black | 17.0 | 19.9 | 22.8 | 18.3 | 21.2 | 24.0 |
| PCV31 | 80 | Black | 6.9 | 8.4 | 9.8 | 7.0 | 8.4 | 9.7 |
| PCV31 | 85 | Black | 3.0 | 5.6 | 7.1 | 3.5 | 5.5 | 7.0 |
| PCV20 | 50 | Non-Black | 6.7 | 15.1 | 31.4 | 8.2 | 18.1 | 37.0 |
| PCV20 | 55 | Non-Black | 8.5 | 16.7 | 30.3 | 10.4 | 20.2 | 36.7 |
| PCV20 | 60 | Non-Black | 11.0 | 19.4 | 31.7 | 13.8 | 23.4 | 37.3 |
| PCV20 | 65 | Non-Black | 12.5 | 19.5 | 28.3 | 15.5 | 23.6 | 33.8 |
| PCV20 | 70 | Non-Black | 12.7 | 17.7 | 23.8 | 15.0 | 20.8 | 27.4 |
| PCV20 | 75 | Non-Black | 10.3 | 13.9 | 18.1 | 11.1 | 14.8 | 19.2 |
| PCV20 | 80 | Non-Black | 4.6 | 6.5 | 8.7 | 4.7 | 6.5 | 8.7 |
| PCV20 | 85 | Non-Black | 2.5 | 4.9 | 7.7 | 2.7 | 4.9 | 7.4 |
| PCV21 | 50 | Non-Black | 9.0 | 19.2 | 38.9 | 10.8 | 22.8 | 46.8 |
| PCV21 | 55 | Non-Black | 11.7 | 21.9 | 39.5 | 14.1 | 26.3 | 46.4 |
| PCV21 | 60 | Non-Black | 15.9 | 26.3 | 41.8 | 19.2 | 31.5 | 49.1 |
| PCV21 | 65 | Non-Black | 18.0 | 27.0 | 39.1 | 21.7 | 32.8 | 46.2 |
| PCV21 | 70 | Non-Black | 18.5 | 25.3 | 33.2 | 21.9 | 29.6 | 38.3 |
| PCV21 | 75 | Non-Black | 15.6 | 20.5 | 25.9 | 16.5 | 21.7 | 26.9 |
| PCV21 | 80 | Non-Black | 7.2 | 9.8 | 12.8 | 7.4 | 9.8 | 12.7 |
| PCV21 | 85 | Non-Black | 4.0 | 7.7 | 11.6 | 4.4 | 7.6 | 11.4 |
| PCV31 | 50 | Non-Black | 11.1 | 23.4 | 47.2 | 13.2 | 27.9 | 56.6 |
| PCV31 | 55 | Non-Black | 14.3 | 26.3 | 46.5 | 16.6 | 31.5 | 56.4 |
| PCV31 | 60 | Non-Black | 19.1 | 31.3 | 48.9 | 22.2 | 37.6 | 59.6 |
| PCV31 | 65 | Non-Black | 22.0 | 31.9 | 44.6 | 25.8 | 38.5 | 54.9 |
| PCV31 | 70 | Non-Black | 22.2 | 29.5 | 37.4 | 26.0 | 34.7 | 44.5 |
| PCV31 | 75 | Non-Black | 18.6 | 23.7 | 29.3 | 20.0 | 25.2 | 31.2 |
| PCV31 | 80 | Non-Black | 8.4 | 11.2 | 14.4 | 8.5 | 11.3 | 14.6 |
| PCV31 | 85 | Non-Black | 4.4 | 8.7 | 12.9 | 4.9 | 8.7 | 13.1 |

| Supplementary Table 2. Number needed to vaccine by each PCV to prevent a case of IPD | | | | | | | | |
|---|---|---|---|---|---|---|---|---|
| | | | **Fast Waning VE** | | | **Slow Waning VE** | | |
| **Vaccine** | **Vac.age** | **Race group** | **2.50%** | **50%** | **97.50%** | **2.50%** | **50%** | **97.50%** |
| PCV20 | 50 | Black | 784 | 1365 | 2000 | 655 | 1142 | 1663 |
| PCV20 | 55 | Black | 752 | 1272 | 1685 | 629 | 1065 | 1407 |
| PCV20 | 60 | Black | 728 | 1185 | 1505 | 601 | 992 | 1242 |
| PCV20 | 65 | Black | 695 | 1102 | 1396 | 570 | 918 | 1137 |
| PCV20 | 70 | Black | 656 | 1018 | 1320 | 559 | 882 | 1115 |
| PCV20 | 75 | Black | 654 | 1032 | 1360 | 618 | 973 | 1272 |
| PCV20 | 80 | Black | 870 | 1445 | 2008 | 871 | 1440 | 1978 |
| PCV20 | 85 | Black | 4022 | 7652 | 15337 | 4017 | 7615 | 13238 |
| PCV21 | 50 | Black | 534 | 1002 | 1590 | 434 | 838 | 1310 |
| PCV21 | 55 | Black | 511 | 916 | 1330 | 420 | 763 | 1096 |
| PCV21 | 60 | Black | 487 | 838 | 1140 | 399 | 700 | 941 |
| PCV21 | 65 | Black | 459 | 770 | 1020 | 381 | 637 | 838 |
| PCV21 | 70 | Black | 431 | 702 | 926 | 371 | 597 | 781 |
| PCV21 | 75 | Black | 427 | 695 | 928 | 401 | 647 | 864 |
| PCV21 | 80 | Black | 568 | 938 | 1322 | 560 | 936 | 1309 |
| PCV21 | 85 | Black | 2542 | 4904 | 9728 | 2595 | 4863 | 8691 |
| PCV31 | 50 | Black | 436 | 834 | 1308 | 371 | 700 | 1096 |
| PCV31 | 55 | Black | 417 | 772 | 1083 | 353 | 645 | 915 |
| PCV31 | 60 | Black | 403 | 712 | 945 | 342 | 594 | 780 |
| PCV31 | 65 | Black | 388 | 657 | 859 | 324 | 545 | 695 |
| PCV31 | 70 | Black | 367 | 604 | 788 | 317 | 519 | 663 |
| PCV31 | 75 | Black | 376 | 594 | 802 | 354 | 564 | 739 |
| PCV31 | 80 | Black | 501 | 818 | 1155 | 494 | 813 | 1152 |
| PCV31 | 85 | Black | 2287 | 4351 | 8596 | 2308 | 4311 | 7635 |
| PCV20 | 50 | Non-Black | 1515 | 3454 | 7942 | 1268 | 2903 | 6623 |
| PCV20 | 55 | Non-Black | 1507 | 3051 | 6052 | 1265 | 2549 | 5059 |
| PCV20 | 60 | Non-Black | 1462 | 2711 | 4801 | 1235 | 2228 | 3998 |
| PCV20 | 65 | Non-Black | 1396 | 2378 | 3857 | 1160 | 1968 | 3189 |
| PCV20 | 70 | Non-Black | 1314 | 2086 | 3289 | 1116 | 1787 | 2767 |
| PCV20 | 75 | Non-Black | 1288 | 1983 | 3080 | 1222 | 1849 | 2860 |
| PCV20 | 80 | Non-Black | 1681 | 2586 | 4321 | 1681 | 2557 | 4264 |
| PCV20 | 85 | Non-Black | 7340 | 13407 | 28724 | 7306 | 13238 | 27574 |
| PCV21 | 50 | Non-Black | 1116 | 2663 | 6268 | 918 | 2244 | 5255 |
| PCV21 | 55 | Non-Black | 1066 | 2294 | 4667 | 893 | 1920 | 3947 |
| PCV21 | 60 | Non-Black | 1030 | 1968 | 3544 | 846 | 1645 | 3007 |
| PCV21 | 65 | Non-Black | 967 | 1697 | 2793 | 788 | 1410 | 2327 |
| PCV21 | 70 | Non-Black | 882 | 1464 | 2321 | 758 | 1253 | 1965 |
| PCV21 | 75 | Non-Black | 864 | 1340 | 2121 | 821 | 1265 | 1958 |
| PCV21 | 80 | Non-Black | 1097 | 1710 | 2801 | 1094 | 1704 | 2792 |
| PCV21 | 85 | Non-Black | 4658 | 8624 | 18751 | 4738 | 8667 | 17262 |
| PCV31 | 50 | Non-Black | 946 | 2217 | 5134 | 803 | 1864 | 4304 |
| PCV31 | 55 | Non-Black | 950 | 1918 | 3881 | 781 | 1607 | 3234 |
| PCV31 | 60 | Non-Black | 910 | 1658 | 3011 | 747 | 1372 | 2478 |
| PCV31 | 65 | Non-Black | 853 | 1434 | 2374 | 698 | 1186 | 1965 |
| PCV31 | 70 | Non-Black | 776 | 1248 | 1951 | 668 | 1066 | 1665 |
| PCV31 | 75 | Non-Black | 749 | 1170 | 1817 | 718 | 1098 | 1716 |
| PCV31 | 80 | Non-Black | 963 | 1508 | 2438 | 962 | 1504 | 2455 |
| PCV31 | 85 | Non-Black | 4113 | 7643 | 16211 | 4136 | 7522 | 15262 |

Supplementary Table 3. Number needed to vaccine by each PCV to prevent a case of IPD

**Slow waning vaccine effectiveness (VE), Age-independent initial VE, PCV13 first dose and PCV31 second dose**

| D1 | D2 | Race group | 2.50% | 50% | 97.50% | Race group | 2.50% | 50% | 97.50% | Race group | 2.50% | 50% | 97.50% |
|---|---|---|---|---|---|---|---|---|---|---|---|---|---|
| 50 | 55 | All | 837 | 1560 | 2707 | Black | 604 | 708 | 849 | Non-Black | 974 | 1809 | 3561 |
| 50 | 60 | All | 762 | 1451 | 2501 | Black | 510 | 607 | 733 | Non-Black | 887 | 1705 | 3280 |
| 50 | 65 | All | 648 | 1236 | 2182 | Black | 395 | 473 | 573 | Non-Black | 766 | 1474 | 2875 |
| 50 | 70 | All | 540 | 1031 | 1817 | Black | 298 | 357 | 434 | Non-Black | 654 | 1253 | 2412 |
| 50 | 75 | All | 466 | 902 | 1560 | Black | 235 | 278 | 333 | Non-Black | 594 | 1107 | 2136 |
| 50 | 80 | All | 408 | 770 | 1297 | Black | 190 | 222 | 267 | Non-Black | 522 | 941 | 1794 |
| 50 | 85 | All | 2662 | 5535 | 10905 | Black | 1228 | 1440 | 1733 | Non-Black | 3360 | 6863 | 13802 |
| 55 | 60 | All | 892 | 1509 | 2401 | Black | 592 | 681 | 804 | Non-Black | 1032 | 1761 | 3100 |
| 55 | 65 | All | 703 | 1208 | 1911 | Black | 429 | 497 | 589 | Non-Black | 823 | 1430 | 2485 |
| 55 | 70 | All | 556 | 953 | 1523 | Black | 303 | 354 | 420 | Non-Black | 671 | 1151 | 1977 |
| 55 | 75 | All | 471 | 816 | 1294 | Black | 231 | 269 | 318 | Non-Black | 589 | 993 | 1706 |
| 55 | 80 | All | 407 | 700 | 1099 | Black | 186 | 217 | 256 | Non-Black | 515 | 848 | 1474 |
| 55 | 85 | All | 2795 | 5122 | 9070 | Black | 1247 | 1431 | 1696 | Non-Black | 3518 | 6281 | 11429 |
| 60 | 65 | All | 780 | 1216 | 1755 | Black | 497 | 559 | 643 | Non-Black | 912 | 1413 | 2215 |
| 60 | 70 | All | 573 | 904 | 1314 | Black | 331 | 373 | 433 | Non-Black | 689 | 1064 | 1667 |
| 60 | 75 | All | 470 | 725 | 1070 | Black | 234 | 267 | 313 | Non-Black | 572 | 875 | 1355 |
| 60 | 80 | All | 397 | 609 | 885 | Black | 182 | 209 | 244 | Non-Black | 481 | 727 | 1147 |
| 60 | 85 | All | 2739 | 4560 | 7174 | Black | 1243 | 1416 | 1664 | Non-Black | 3414 | 5487 | 9055 |
| 65 | 70 | All | 755 | 1049 | 1413 | Black | 448 | 495 | 555 | Non-Black | 869 | 1223 | 1756 |
| 65 | 75 | All | 577 | 802 | 1101 | Black | 298 | 336 | 388 | Non-Black | 680 | 955 | 1349 |
| 65 | 80 | All | 452 | 635 | 852 | Black | 219 | 247 | 286 | Non-Black | 540 | 755 | 1100 |
| 65 | 85 | All | 3086 | 4631 | 6730 | Black | 1441 | 1627 | 1911 | Non-Black | 3827 | 5526 | 8408 |
| 70 | 75 | All | 849 | 1077 | 1378 | Black | 474 | 520 | 575 | Non-Black | 986 | 1256 | 1637 |
| 70 | 80 | All | 624 | 810 | 1056 | Black | 325 | 366 | 425 | Non-Black | 733 | 949 | 1276 |
| 70 | 85 | All | 4065 | 5549 | 7604 | Black | 2001 | 2249 | 2606 | Non-Black | 4873 | 6526 | 9308 |
| 75 | 80 | All | 1006 | 1257 | 1590 | Black | 592 | 667 | 764 | Non-Black | 1185 | 1444 | 1835 |
| 75 | 85 | All | 6369 | 8175 | 10853 | Black | 3435 | 3901 | 4503 | Non-Black | 7427 | 9516 | 12859 |
| 80 | 85 | All | 14073 | 18756 | 26277 | Black | 8413 | 9971 | 12001 | Non-Black | 16206 | 21612 | 29690 |

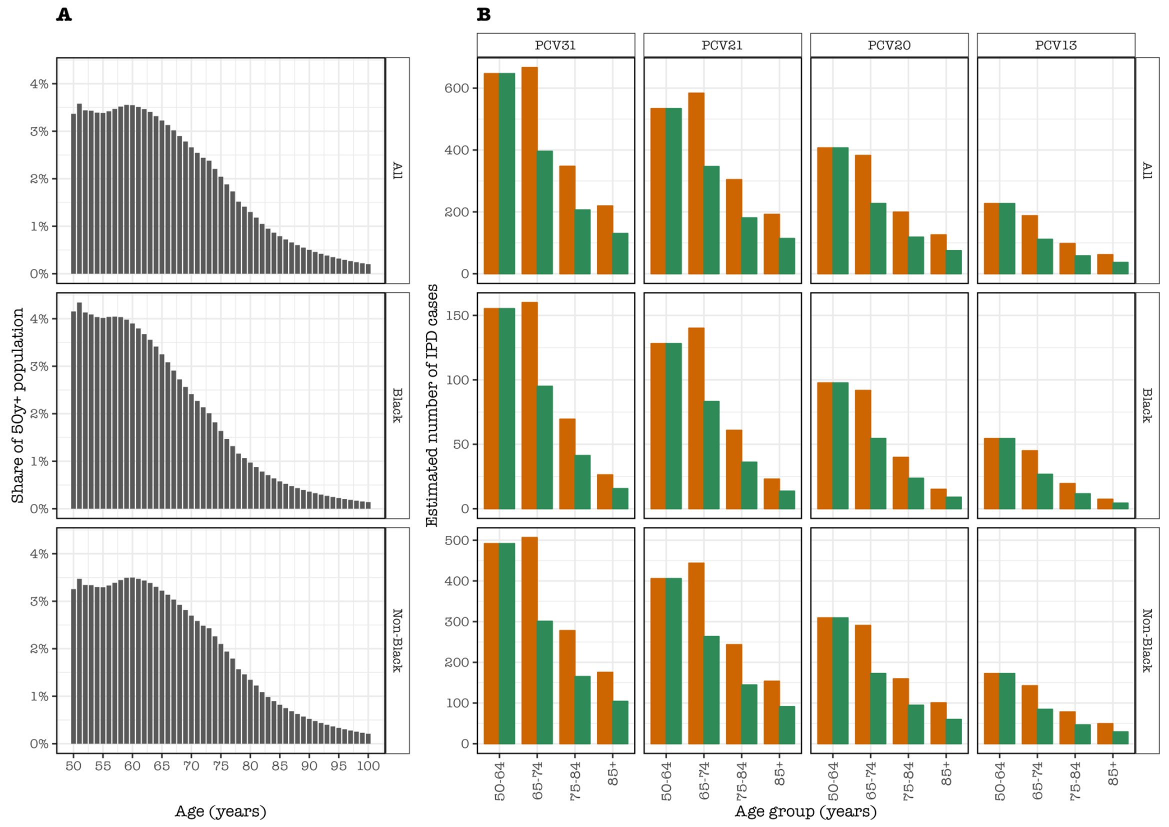


Supplementary Figure 1. (A) Population age distribution by overall population and demographic groups in the catchment area of the Active Bacterial Core surveillance (ABCs) in the United States, smoothed using 5-year moving average. The age distribution among Black population is steeper than non-Black population. (B) The estimated number of IPD cases in each demographic group stratified by serotype group of each pneumococcal conjugate vaccine (PCV) product, age groups and whether PCV immunity among adult population pre-exists (PCV-experienced adult population, Green) or is absent (PCV-naïve adult population, Orange). The estimated number of IPD cases are slightly more under the scenario of PCV-naïve than PCV-experienced population among those aged 65+ years-old.

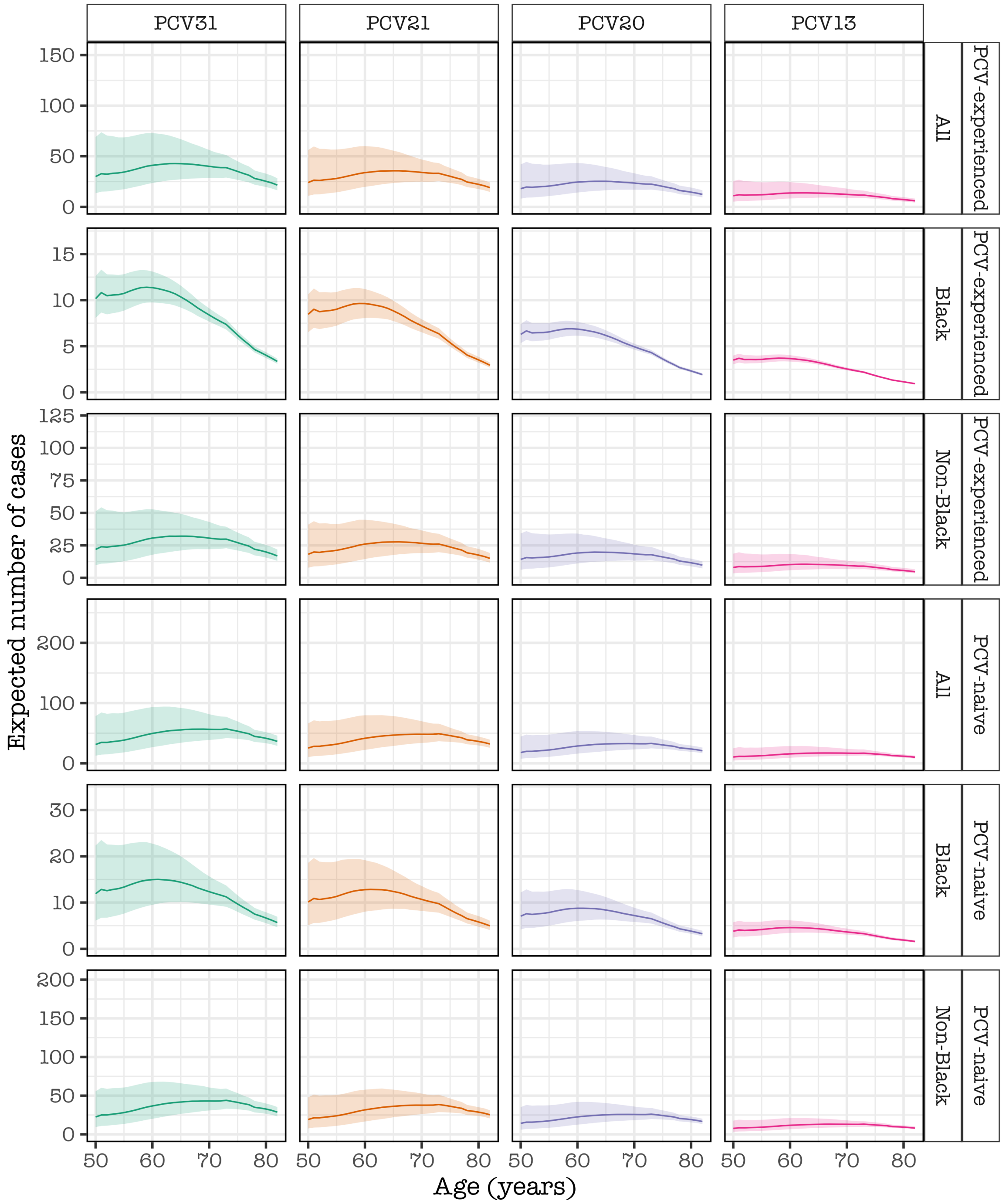


Supplementary Figure 2. The expected number of IPD cases by overall and demographic groups, and serotype group of each pneumococcal conjugate vaccine (PCV) product in single-year age assuming some PCV immunity among adult population pre-exists (PCV-experienced adult population) or is absent (PCV-naïve adult population). The expected number of IPD cases are much higher among younger than older Black adults in constrast to non-Black population.

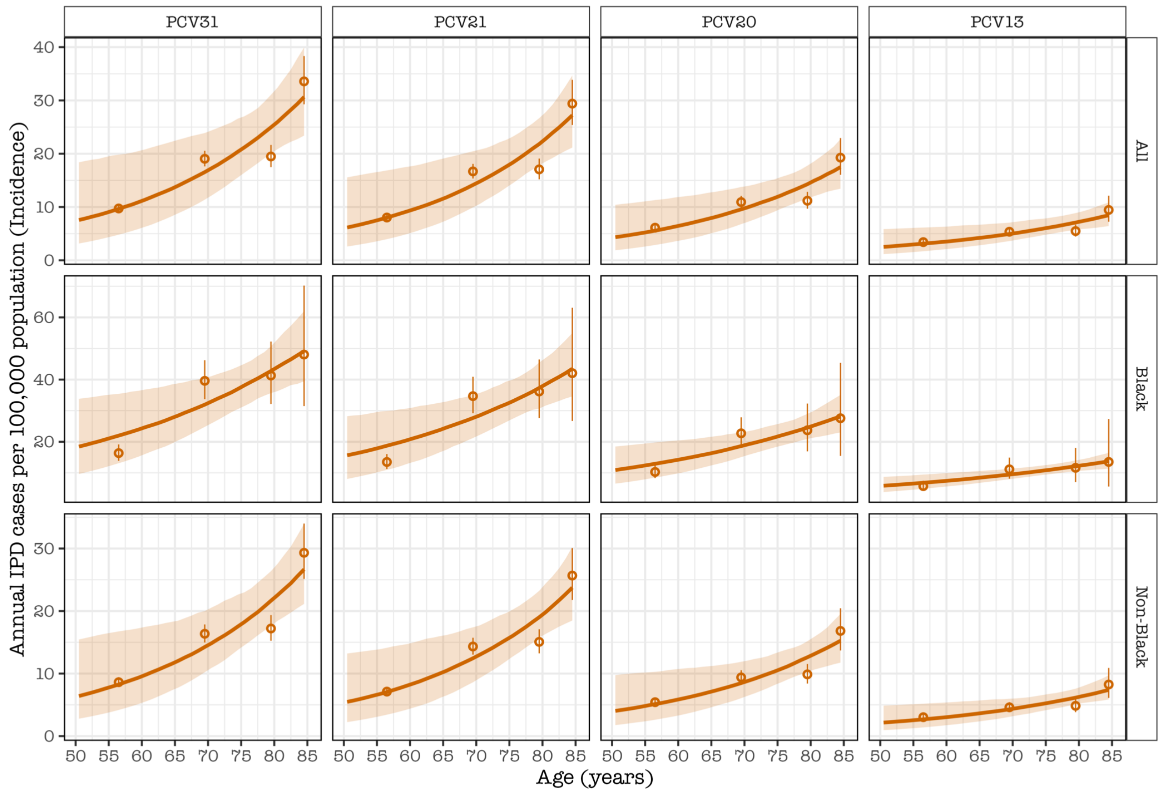


Supplementary Figure 3. The inferred incidence of invasive pneumococcal disease (IPD) by overall and demographic groups, and serotype group of each pneumococcal conjugate vaccine (PCV) product in single-year age **assuming absence of PCV immunity among the adult population** (PCV-naïve adult population) in the United States. The solid line and shaded ribbon refer to mean incidence and 95% confidence intervals (CI) of the mean incidence, and the circles and vertical lines refer to inferred reported IPD cases per 100,000 population and corresponding 95%CI.

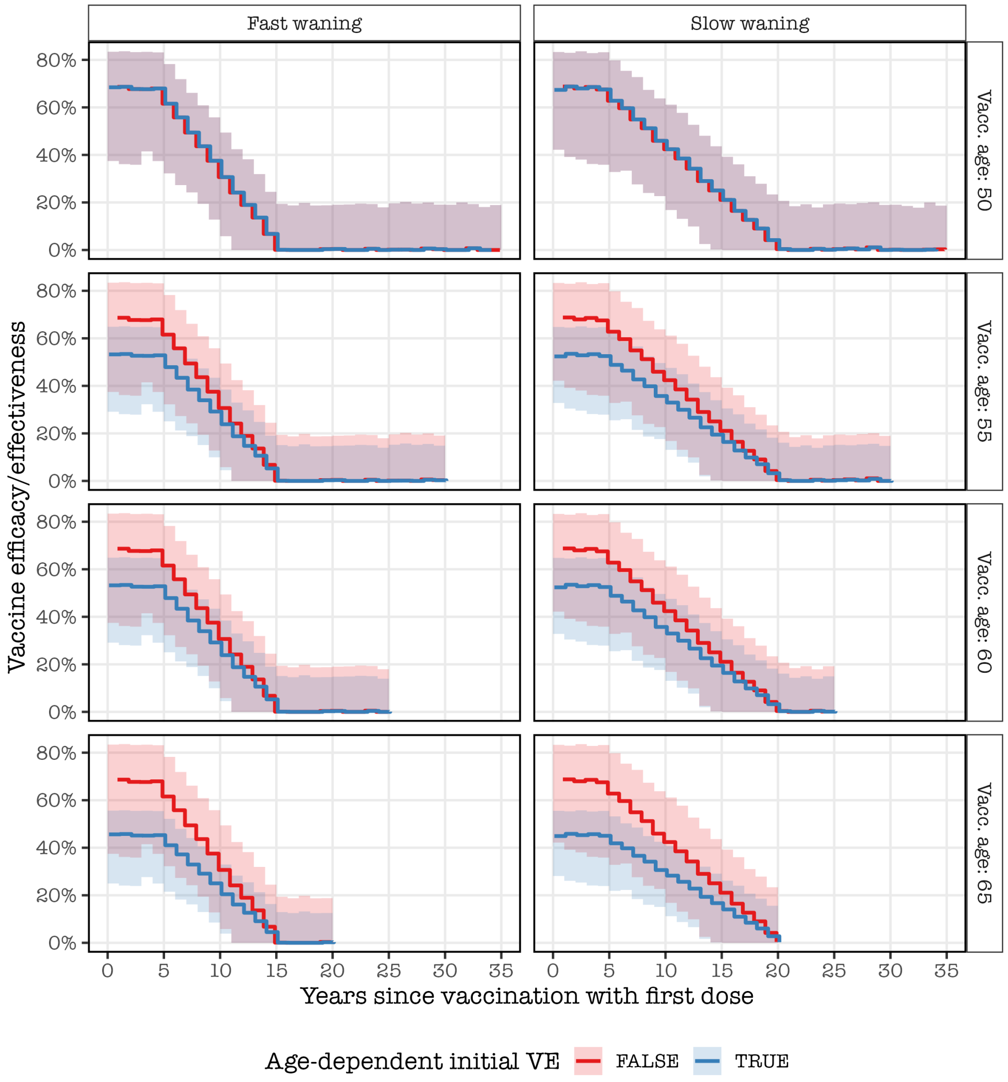


Supplementary Figure 4. A snapshot based on the Centers for Disease Control and Prevention (CDC)'s Advisory Committee on Immunisation Practices (ACIP) assumptions about initial vaccine effectiveness (VE) and waning VE from the time since vaccination stratified by age of vaccination, waning VE categories, and age-dependency of initial VE. The solid line and shaded ribbon refer to mean VE and 95% confidence intervals (CI) of the mean VE. The plot contrasts between fast and slow waning VE, age-dependent, and age-independent initial VE in 5-year intervals of the age of vaccination.

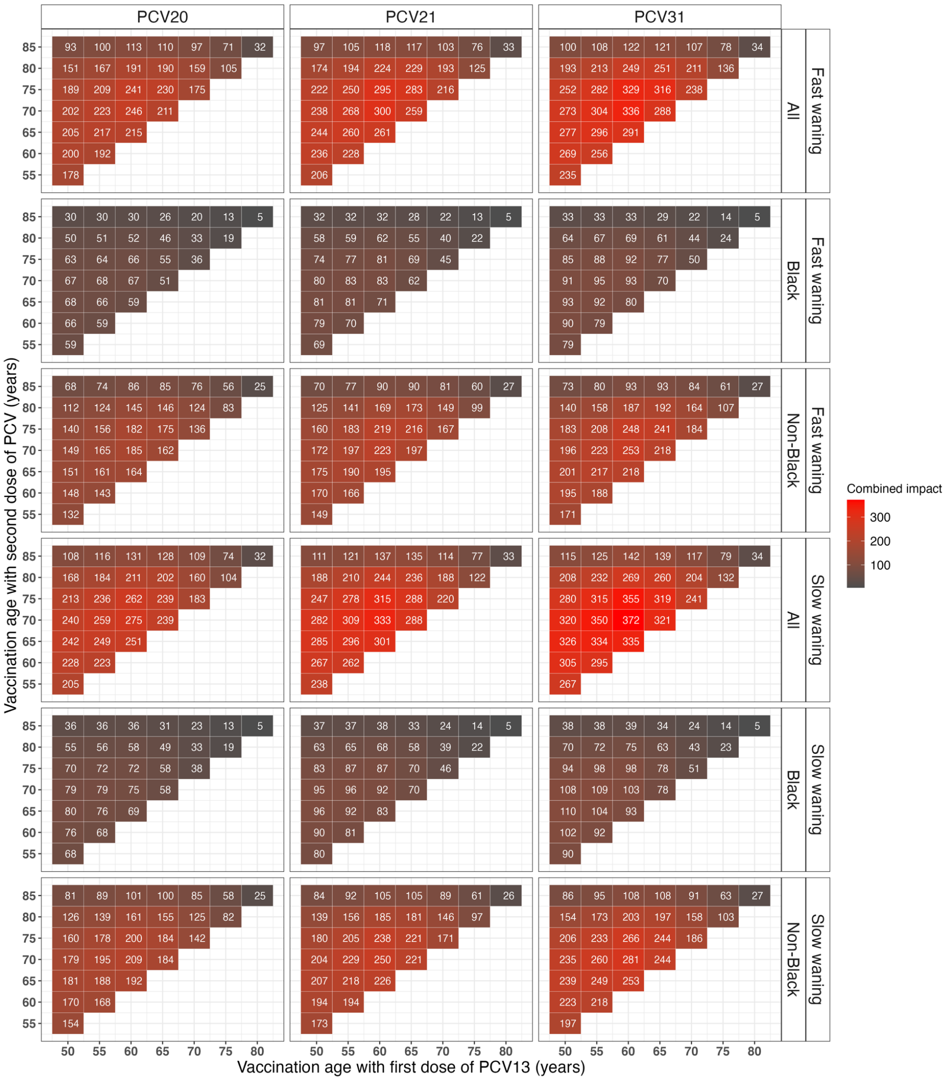


Supplementary Figure 5. The combined predicted number of IPD cases averted for the rest of the age cohort lifetime for administering the first dose of PCV13 at a specific age followed by the second dose of higher-valency PCV at a range of 5-year ages, stratified by pneumococcal conjugate vaccine (PCV) product, demographic group, and waning PCV effectiveness (VE) category, under the scenarios of **ACIP age-independent initial VE, and the presence of prior PCV immunity in adult population** in the United States. Numbers in the heatmap cells show cohort model mean estimates and the color scale indicate whether more or less cases are preventable. The optimal ages of the first and second doses combined is similar among Black and non-Black adults, and is at 5 years of the first dose in late adulthood or at 10 or 15 years of the first dose in early adulthood.

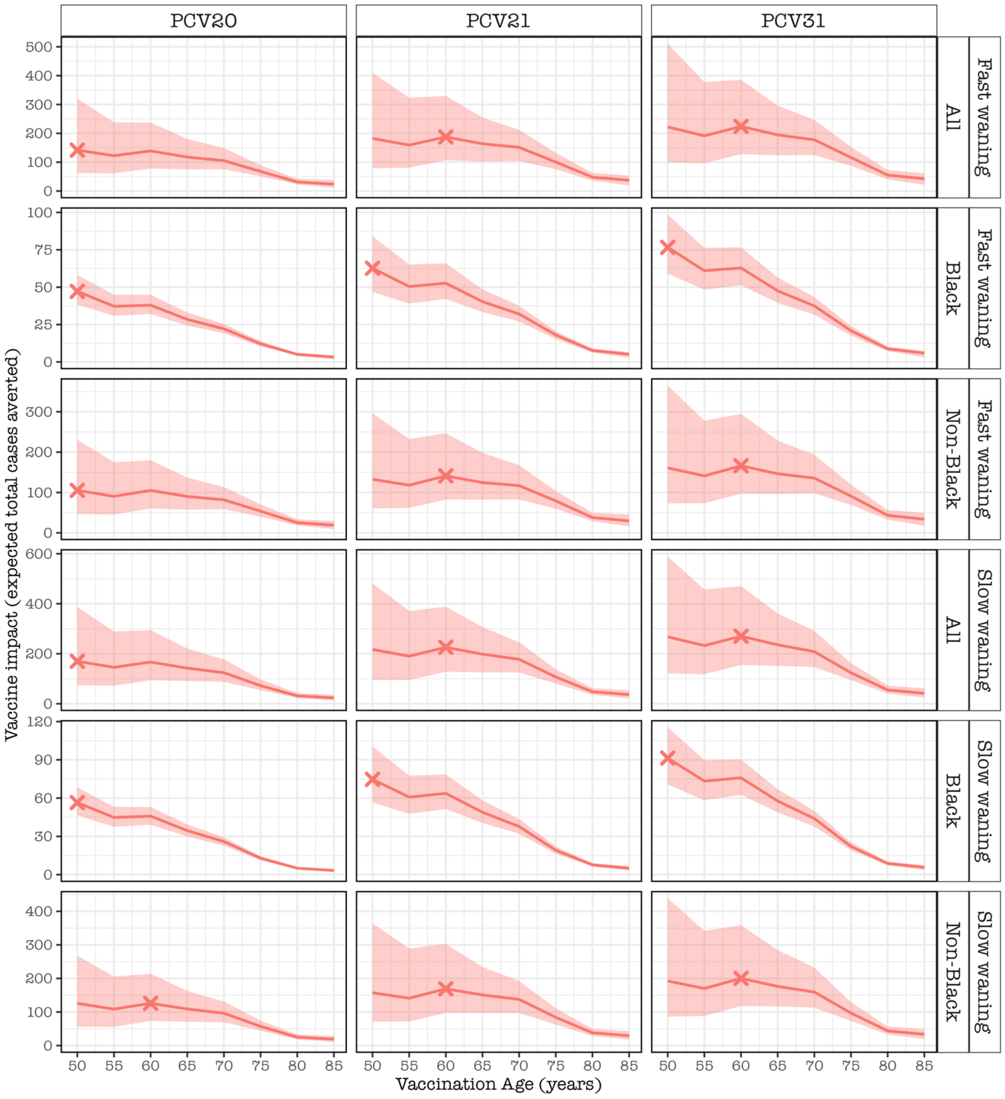


Supplementary Figure 6. The expected number of IPD cases averted for the rest of the age cohort lifetime by vaccinating every older adult in the age cohort, stratified by pneumococcal conjugate vaccine (PCV) product, demographic group, and waning PCV effectiveness (VE) category, under the scenarios of **ACIP age-dependent initial VE and waning VE and presence of prior PCV immunity** in adult population in the United States. The red line and shaded ribbon represent cohort model mean estimates and 95% bootstrap confidence intervals for the mean estimates. The X corresponds to the optimal age for pneumococcal vaccination. Overall, most cases are preventable at 50 years or 60 years; among Black adults at 50 years whereas among non-Black, this is achieved at 60 years.

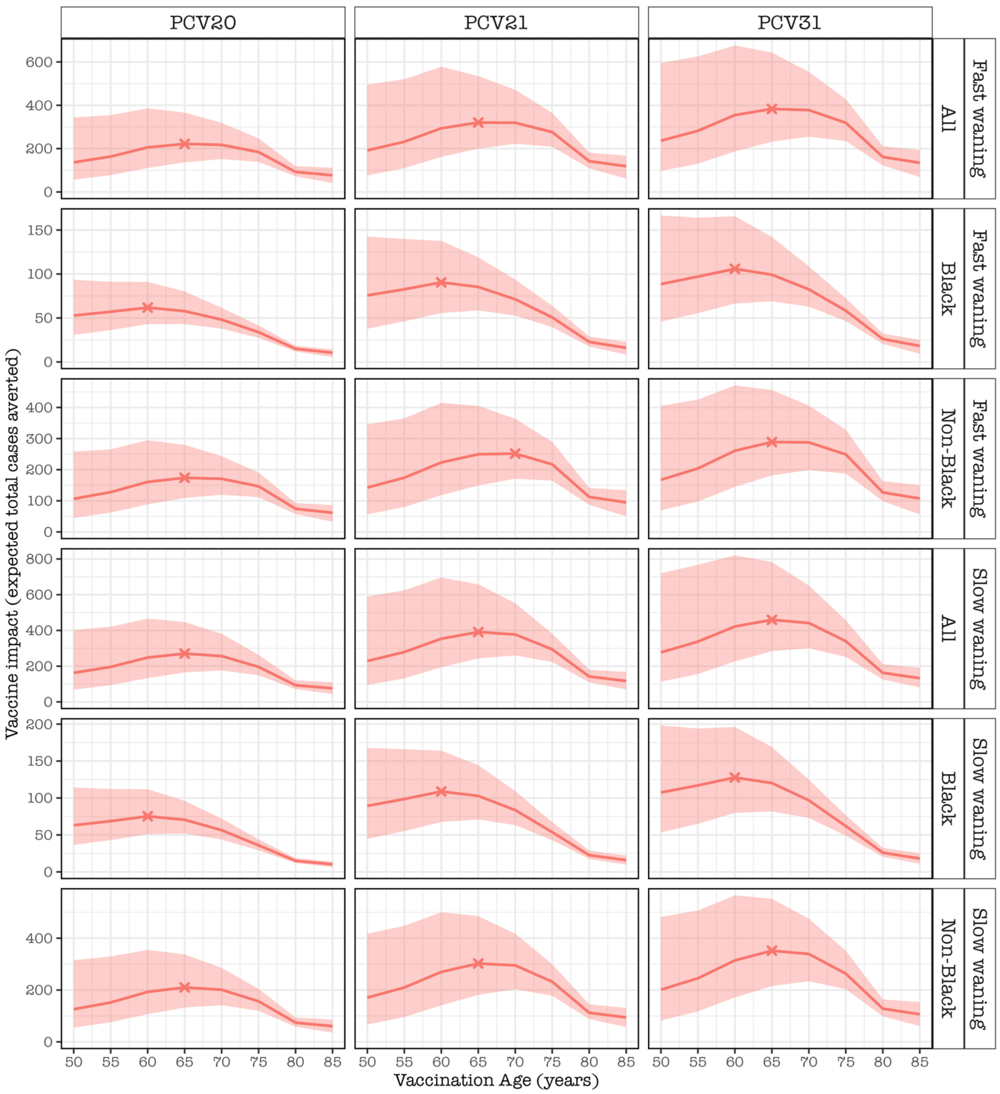


Supplementary Figure 7. The expected number of IPD cases averted for the rest of the age cohort lifetime by vaccinating every older adult in the age cohort, stratified by pneumococcal conjugate vaccine (PCV) product, demographic group, and waning PCV effectiveness (VE) category, under the scenario of **ACIP age-independent initial VE and waning VE and absence of prior PCV immunity** in adult population in the United States. The red line and shaded ribbon represent cohort model mean estimates and 95% bootstrap confidence intervals for the mean estimates. The X corresponds to the optimal age for pneumococcal vaccination. Overall, most cases are preventable at 65 years; among Black adults at 60 years and among non-Black at either 65 years or 70 years (depending on waning VE assumption).

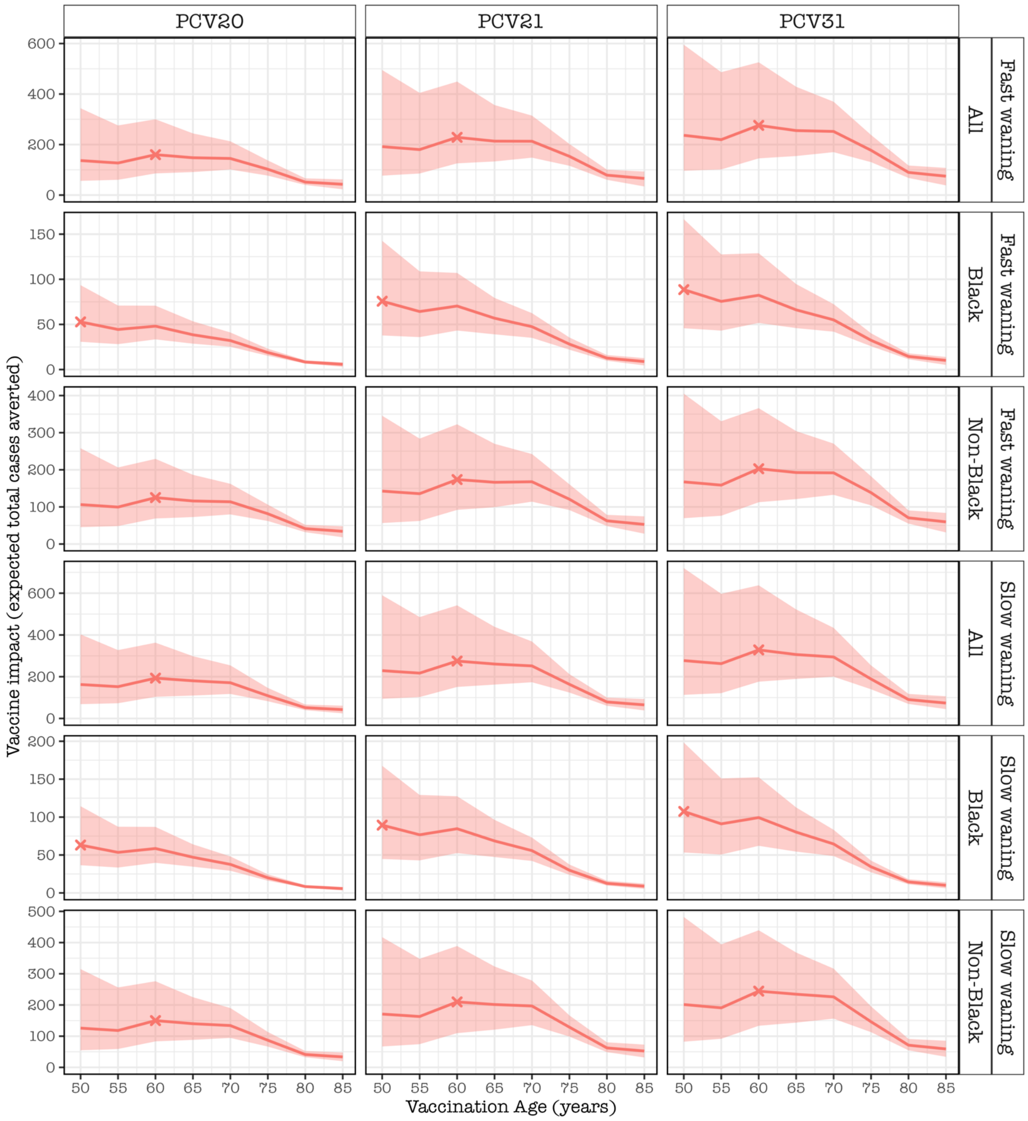


Supplementary Figure 8. The expected number of IPD cases averted for the rest of the age cohort lifetime by vaccinating every older adult in the age cohort, stratified by pneumococcal conjugate vaccine (PCV) product, demographic group and waning PCV effectiveness (VE) category, under the scenario of **ACIP age-dependent initial and waning VE and absence of prior PCV immunity** in adult population in the United States. The red line and shaded ribbon represent cohort model mean estimates and 95% bootstrap confidence intervals for the mean estimates. The X corresponds to the optimal age for pneumococcal vaccination. Overall, most cases are preventable at 60 years; among Black adults at 50 years and among non-Black at 60 years.

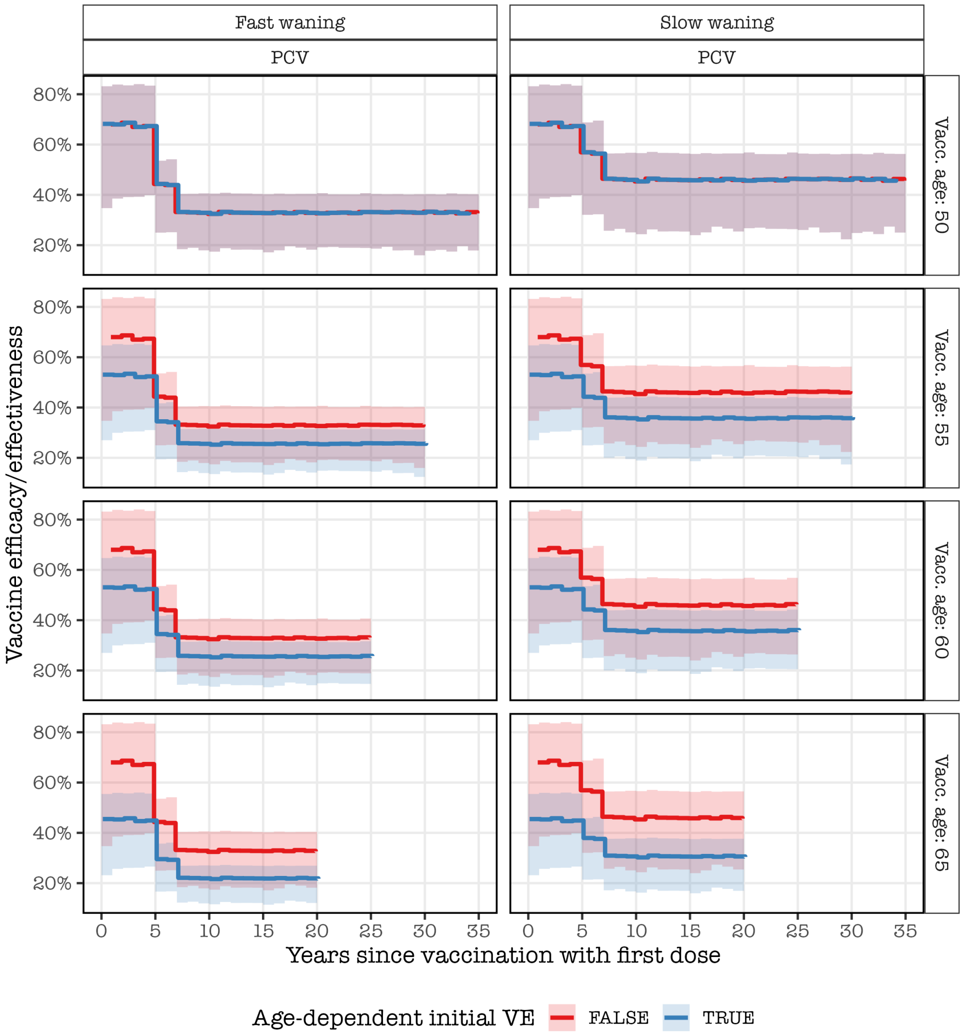


Supplementary Figure 9. Stepwise-based initial vaccine effectiveness (VE) and waning VE from the time since vaccination **based on stepwise decay model** described above in Supplementary Text 2, stratified by age of vaccination, waning VE categories and age-dependency of initial VE. The solid line and shaded ribbon refer to mean VE and 95% confidence intervals (CI) of the mean VE. The plot contrasts between fast and slow waning VE, age-dependent and age-independent initial VE in 5-year intervals of age of vaccination.

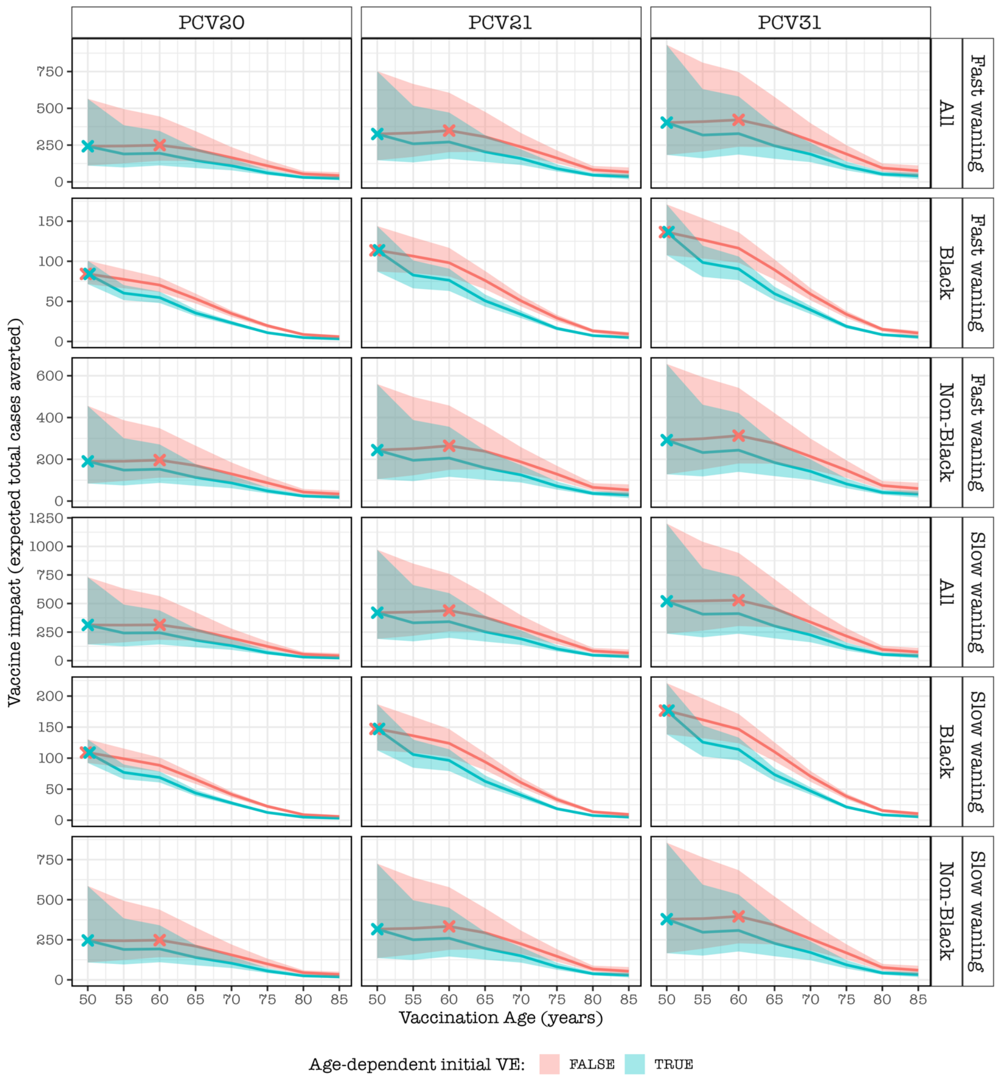


Supplementary Figure 10. The expected number of IPD cases averted for the rest of the age cohort lifetime by vaccinating every older adult in the age cohort, stratified by pneumococcal conjugate vaccine (PCV) product, demographic group and waning PCV effectiveness (VE) category, **under the scenarios of stepwise waning VE and presence of prior PCV immunity** in adult population in the United States. The line and shaded ribbon represent cohort model mean estimates and 95% bootstrap confidence intervals for the mean estimates. The X corresponds to the optimal age for pneumococcal vaccination. Most cases are preventable at 50 years regardless of demographics if age-dependent initial VE is assumed. However, if age-independent initial VE is assumed, most cases are preventable at 50 years among Black adults, and at 60y among non-Black adults and overall population.

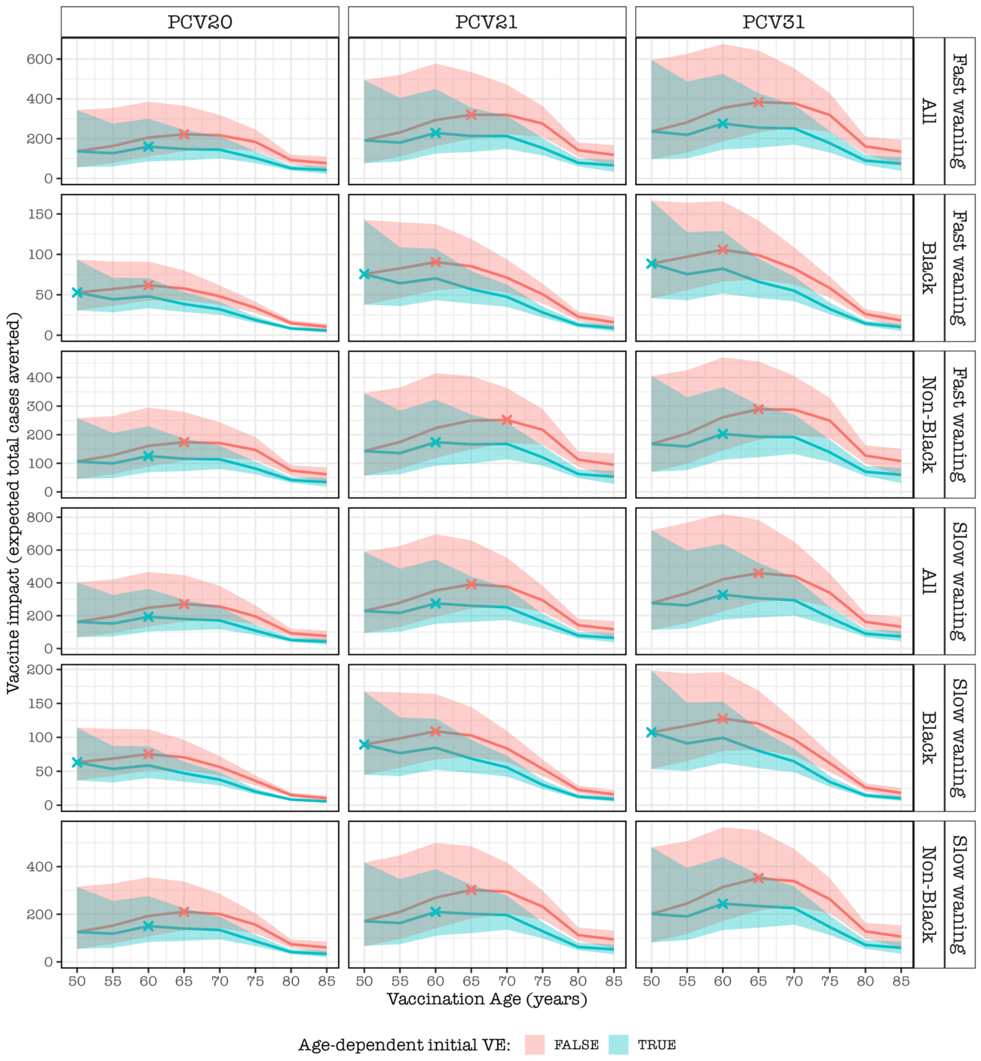


Supplementary Figure 11. The expected number of IPD cases averted for the rest of the age cohort lifetime by vaccinating every older adult in the age cohort, stratified by pneumococcal conjugate vaccine (PCV) product, demographic group and waning PCV effectiveness (VE) category, **under the scenarios of stepwise waning VE and absence of prior PCV immunity** in adult population in the United States. The line and shaded ribbon represent cohort model mean estimates and 95% bootstrap confidence intervals for the mean estimates. The X corresponds to the optimal age for pneumococcal vaccination. Among Black adults, most cases are preventable at 50 years if age-dependent initial VE or at 60y if age-independent initial VE is assumed. Among Non-Black adults or overall population, most cases are preventable at 60 years if age-dependent initial VE or at 65y if age-independent initial VE is assumed.

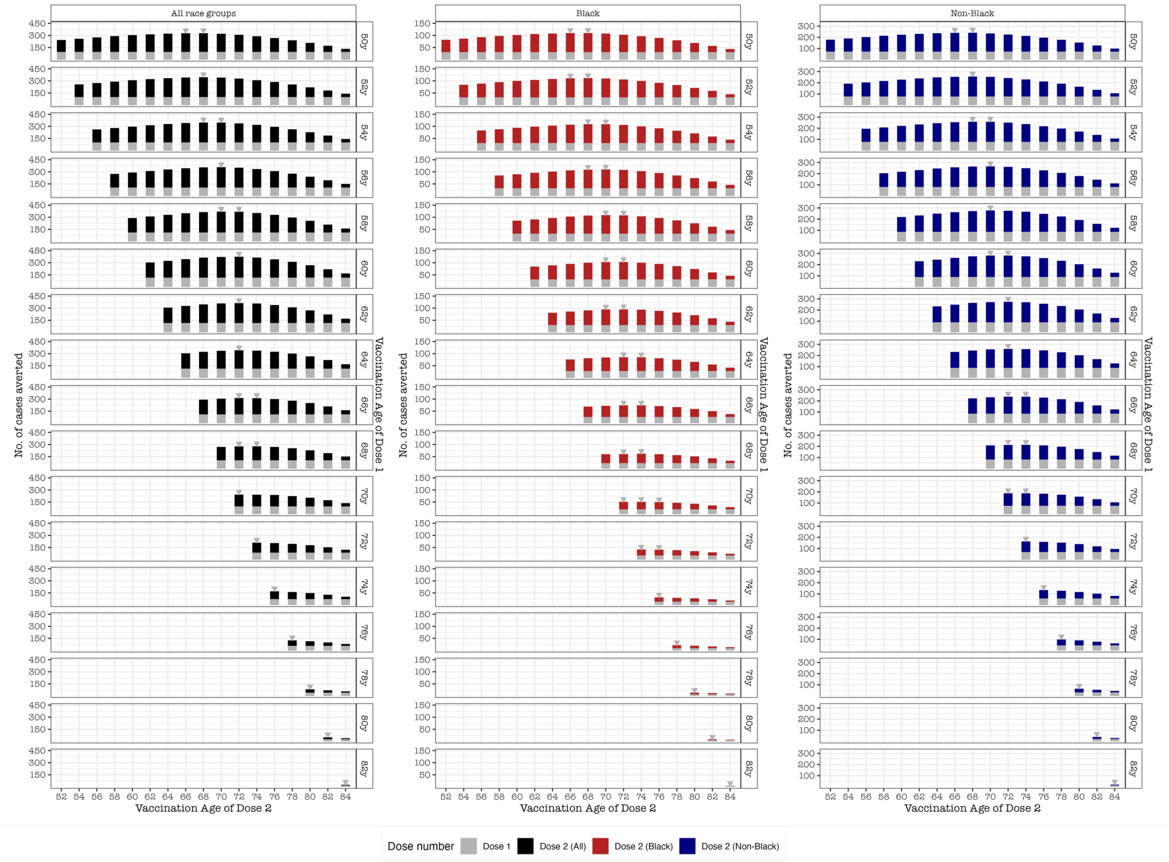


Supplementary Figure 12. The predicted number of IPD cases averted for the rest of the age cohort lifetime, **under high-resolution vaccination age intervals**, for administering the first dose of PCV13 at a specific age (right y-axis) followed by the second dose of higher-valency PCV at any of the ages (x-axis), stratified by pneumococcal conjugate vaccine (PCV) product, demographic group and waning PCV effectiveness (VE) category, under the scenarios of ACIP initial and waning VE and presence of prior PCV immunity in adult population in the United States. The bars show cohort model mean estimates and the red color indicate second dose Black vaccinees and blue color indicate non-Black vaccinees. The optimal ages of the first and second doses are similar regardless of demographics. The second dose should be given as soon as possible if receipt of first dose was in late adulthood (70y+) or the second dose should be given at an age corresponding to initial dose, which largely reflects decreasing years to second dose with increasing age from 50 to 69 years-old.
